# Geographical concentration of COVID-19 cases by social determinants of health in 16 large metropolitan areas in Canada – a cross-sectional study

**DOI:** 10.1101/2021.07.23.21261039

**Authors:** Yiqing Xia, Huiting Ma, Gary Moloney, Héctor A. Velásquez García, Monica Sirski, Naveed Z. Janjua, David Vickers, Tyler Williamson, Alan Katz, Kristy Yu, Rafal Kustra, David L Buckeridge, Marc Brisson, Stefan D Baral, Sharmistha Mishra, Mathieu Maheu-Giroux

**Author notes:** **Corresponding author** Sharmistha Mishra, Mailing address: Room 315, MAP Centre for Urban Health Solutions, Li Ka Shing Knowledge Institute, 209 Victoria St., Toronto, ON Canada M5B 1T8. contributed equally (Yiqing Xia, Huiting Ma). **Conflict of interest** MM-G report contractual agreements with the *Institut national de santé publique du Québec (INSPQ)* and the *Institut d’excellence en santé et en services sociaux* (INESSS).

## Abstract

**Background:** There is a growing recognition that strategies to reduce SARS-CoV-2 transmission should be responsive to local transmission dynamics. Studies have revealed inequalities along social determinants of health, but little investigation was conducted surrounding geographic concentration within cities. We quantified social determinants of geographic concentration of COVID-19 cases across sixteen census metropolitan areas (CMA) in four Canadian provinces.

**Methods:** We used surveillance data on confirmed COVID-19 cases at the level of dissemination area. Gini (co-Gini) coefficients were calculated by CMA based on the proportion of the population in ranks of diagnosed cases and each social determinant using census data (income, education, visible minority, recent immigration, suitable housing, and essential workers) and the corresponding share of cases. Heterogeneity was visualized using Lorenz (concentration) curves.

**Results:** Geographic concentration was observed in all CMAs (half of the cumulative cases were concentrated among 21-35% of each city’s population): with the greatest geographic heterogeneity in Ontario CMAs (Gini coefficients, 0.32-0.47), followed by British Columbia (0.23-0.36), Manitoba (0.32), and Québec (0.28-0.37). Cases were disproportionately concentrated in areas with lower income, education attainment, and suitable housing; and higher proportion of visible minorities, recent immigrants, and essential workers. Although a consistent feature across CMAs was concentration by proportion visible minorities, the magnitude of concentration by social determinants varied across CMAs.

**Interpretation:** The feature of geographical concentration of COVID-19 cases was consistent across CMAs, but the pattern by social determinants varied. Geographically-prioritized allocation of resources and services should be tailored to the local drivers of inequalities in transmission in response to SARS-CoV-2’s resurgence.

## Introduction

The COVID-19 epidemics in Canada have varied in size and trajectory across provinces and their large cities (1, 2). At the national-level (3) and within provinces (4, 5), there has been a disproportionate burden of confirmed cases, and thus severe outcomes, among socially and economically marginalized communities (6). Social determinants of health refer to non-medical factors influencing health outcomes whereas structural determinants encompass cultural norms, policies, and institutions that generate social stratification and determine socio-economic position (4, 5). In Canada and elsewhere, data have consistently highlighted the importance of determinants such as household size and density, work in essential services, and proxies for structural racism in the relative risk of COVID-19 (6-14).

Understanding the factors leading to geographical patterning of transmission within cities can help identify the populations, and specifically the contexts, with the greatest risks; analyses which enable better allocation of resources, tailoring of policies, and implementation of context-specific strategies to more effectively and efficiently curb local transmission (15). To date, few studies have quantified and compared the geographical concentration of COVID-19 cases by social determinants across Canada, and the extent to which the magnitude of inequalities might vary between cities and provinces. We therefore sought to quantify and compare the magnitude of geographical concentration of cases by area-level social determinants of health across 16 metropolitan areas in four Canadian provinces: British Columbia, Manitoba, Ontario, and Québec. Together these provinces accounted for 79% of cases by July 8th, 2021.

## Methods

### Study design and study population

We conducted a cross-sectional study using surveillance data from four provinces, over the January 23, 2021 (report date of the first documented case in Canada) to February 28, 2021 period. We restricted analyses to the country’s largest census metropolitan areas (CMA) that accounted for more than 80% of diagnosed COVID-19 cases in each province; except for Manitoba where only Winnipeg is qualified as CMA by census definition(16).

Table 1 summarizes the characteristics of the CMAs. Due to the unique context of transmission in long-term care homes, we excluded cases among their residents to focus on transmission dynamics in the wider community. The unit of analysis was the dissemination area (DA), which is the smallest standard geographic unit with census information, representing between 400-700 residents (17).

**Table 1.** Characteristics of census metropolitan areas (CMA) and dissemination areas (DA) included in the study (19).

| Census Metropolitan Areas | Population | Cases (N) | Pop with 50% cases<sup>a</sup> (%) | DAs (N) | DA with no/missing cases (%) |
| --- | --- | --- | --- | --- | --- |
| **British Columbia** |  |  |  |  |  |
| Vancouver | 2,454,378 | 54,222 | 25.8% | 3,425 | 2.7% |
| Kelowna | 184,190 | 2,865 | 34.7% | 239 | 3.8% |
| Abbotsford-Mission | 180,230 | 5,622 | 27.5% | 263 | 2.3% |
| **Manitoba** |  |  |  |  |  |
| Winnipeg | 777,496 | 15,089 | 28.5% | 1,224 | 8.3% |
| **Ontario** |  |  |  |  |  |
| Toronto | 5,927,779 | 187,764 | 29.1% | 7,522 | 0.2% |
| Ottawa-Gatineau (Ontario part) | 991,726 | 13,975 | 21.2% | 1,456 | 1.7% |
| Hamilton | 747,545 | 12,490 | 26.1% | 1,199 | 0.8% |
| Kitchener-Cambridge-Waterloo | 523,894 | 9,598 | 29.6% | 736 | 0.4% |
| St. Catharines-Niagara | 406,074 | 6,835 | 23.6% | 678 | 1.6% |
| Windsor | 329,144 | 8,498 | 29.7% | 548 | 8.0% |
| **Québec** |  |  |  |  |  |
| Montréal | 4,098,927 | 175,111 | 29.3% | 6,469 | 6.5% |
| Québec City | 800,296 | 22,219 | 30.3% | 1,291 | 5.6% |
| Ottawa-Gatineau (Québec part) | 332,057 | 5,337 | 33.1% | 491 | 4.9% |
| Sherbrooke | 212,105 | 4,572 | 29.2% | 327 | 6.4% |
| Saguenay | 160,980 | 5,056 | 28.2% | 295 | 6.1% |
| Trois-Rivières | 156,042 | 3,633 | 33.5% | 272 | 4.8% |

<sup>a</sup>Pop with 50% cases = Percentage of population that accounted for 50% of the total cases.
<sup>b</sup>Pop = population size.
<sup>c</sup>Income = after-tax income per person equivalent
<sup>d</sup>Diploma = proportion without certificate, degree, or diploma.
<sup>e</sup>Visible minority = proportion visible minority.
<sup>f</sup>Recent immigration = proportion of recent immigration.
<sup>g</sup>Suitable housing = proportion with suitable housing.
<sup>h</sup>Essential workers = proportion essential worker

### Data sources

Individual-level data from provincial surveillance databases were used to calculate the number of SARS-CoV-2 cases per DA. In British Columbia, confirmed cases are recorded in case line list integrated in the *Public Health Reporting Data Warehouse*. In Manitoba, the COVID-19 surveillance data and contact investigation information were requested through the *Manitoba Population Research Data Repository* (18). In Ontario, data on laboratory-confirmed cases were recorded in the *case contact management solutions*. In Québec, confirmed cases were recorded in the *Trajectoire de santé publique* database. For each confirmed case, basic sociodemographic information was collected (i.e., address) in addition to epidemiological characteristic such as date of case report and living environment (e.g., long-term care facility). Cases were assigned to a DA according to the residential address using the *Postal Code Conversion File* (19) for all provinces.

Data describing DA-level social determinants of health, with the exception of income, were extracted from the latest available Canadian census data (2016) (20). The after-tax income per person equivalent ranking across DAs was obtained from the *Postal Code Conversion File Plus Version 7A/7D* for each provinces (21).

### Measures

We defined COVID-19 cases as laboratory-confirmed cases (all provinces). For Québec we also included cases confirmed by epidemiological link (individual with COVID-19 symptoms without other apparent cause that had a close contact with a laboratory-confirmed case (22)) due to lack of testing capacity during the first wave in February – April 2020. We considered the following measures of social determinants of SARS-CoV-2 transmission (23): 1) socio-demographic indices (after-tax income per-person equivalent, proportion population without certificate, diploma or degree, proportion visible minority, proportion recent immigration) (14, 15, 24); 2) dwelling-related indicators (proportion with suitable housing) (6, 10) and, 3) occupation-related variables (proportion working in essential services: health, trades and transport and equipment operation, sales and services, manufacturing and utilities, resources, agriculture and production) (13). Determinants were ranked from the highest value to the lowest and grouped into ten deciles. *Table S1* details the definitions of each variable.

### Analyses

The cumulative numbers of confirmed COVID-19 cases were aggregated to the DA-level, along with population denominators, and social determinants. First, we quantified the magnitude of overall geographical heterogeneity within each CMA using Gini coefficients and crude Lorenz curves. Second, we quantified the extent to which cases were concentrated by each social determinant using co-Gini coefficients and concentration curves. To generate the curves, we plotted the cumulative share of CMA’s population ranked by number of cases or each social determinant on the x-axis and the corresponding cumulative proportion of cases on the y-axis. The Lorenz (concentration) curves depict a diagonal line of equality, and the further the data deviate from the diagonal, the higher the variability (or greater inequality/concentration) in cases across the population. The Gini and co-Gini coefficients were calculated as twice the area between the Lorenz (concentration) curve and the line of equality. (25). Values closer to 1 reflect greater inequality while values closer to 0 represents uniform distributions (26).

Data management and analyses were conducted by each provincial team separately using standardized protocols and a shared code base. Aggregated results were shared across provincial teams as per the data privacy requirements of each province. All analyses were conducted using R statistical software (27).

### Ethics approval

Ethics approvals were obtained from the *Research Ethics Board* of University of British Columbia in British Columbia (H20-02097), the *Health Research Ethics Board* of University of Manitoba (HS24140 (H2020:352)) and the *Health Information Privacy Committee* of the Government of Manitoba (No. 2020/2021-32) in Manitoba, the *Health Sciences Research Ethics Board* of University of Toronto (no. 39253) in Ontario, and the *Institutional Review Board* of McGill University in Québec (A06-M52-20B).

## Results

During the study period, 63,266 (British Columbia), 15,089 (Manitoba), 239,160 (Ontario), and 224,377 (Québec) cases were recorded in the 16 CMAs included in the study. These 16 CMAs accounted for 81%, 57%, 83% and 80% of all confirmed cases in each province, respectively. Less than 9% of the DAs recorded zero cases during the study period (**Table 1**).

### Magnitude of overall heterogeneity between cities

Half of the cumulative COVID-19 cases were diagnosed among approximately 21-35% of the population in each CMAs (**Figure 1, Table 1**). CMAs in Ontario exhibited the greatest heterogeneity (Gini coefficients: 0.32-0.47), followed by British Columbia (Gini coefficients: 0.23-0.36), Manitoba (Gini coefficient: 0.31) and then Québec (Gini coefficients: 0.28-0.37). The magnitude of heterogeneity varied within provinces as well. The largest and smallest Gini coefficients were observed, respectively, in Vancouver and Kelowna in British Columbia; St. Catharines–Niagara and Hamilton in Ontario; and Saguenay and Trois-Rivières in Québec. Lorenz curves and Gini coefficients for each CMA can be found in *Figure S1*.

**Figure 1.**
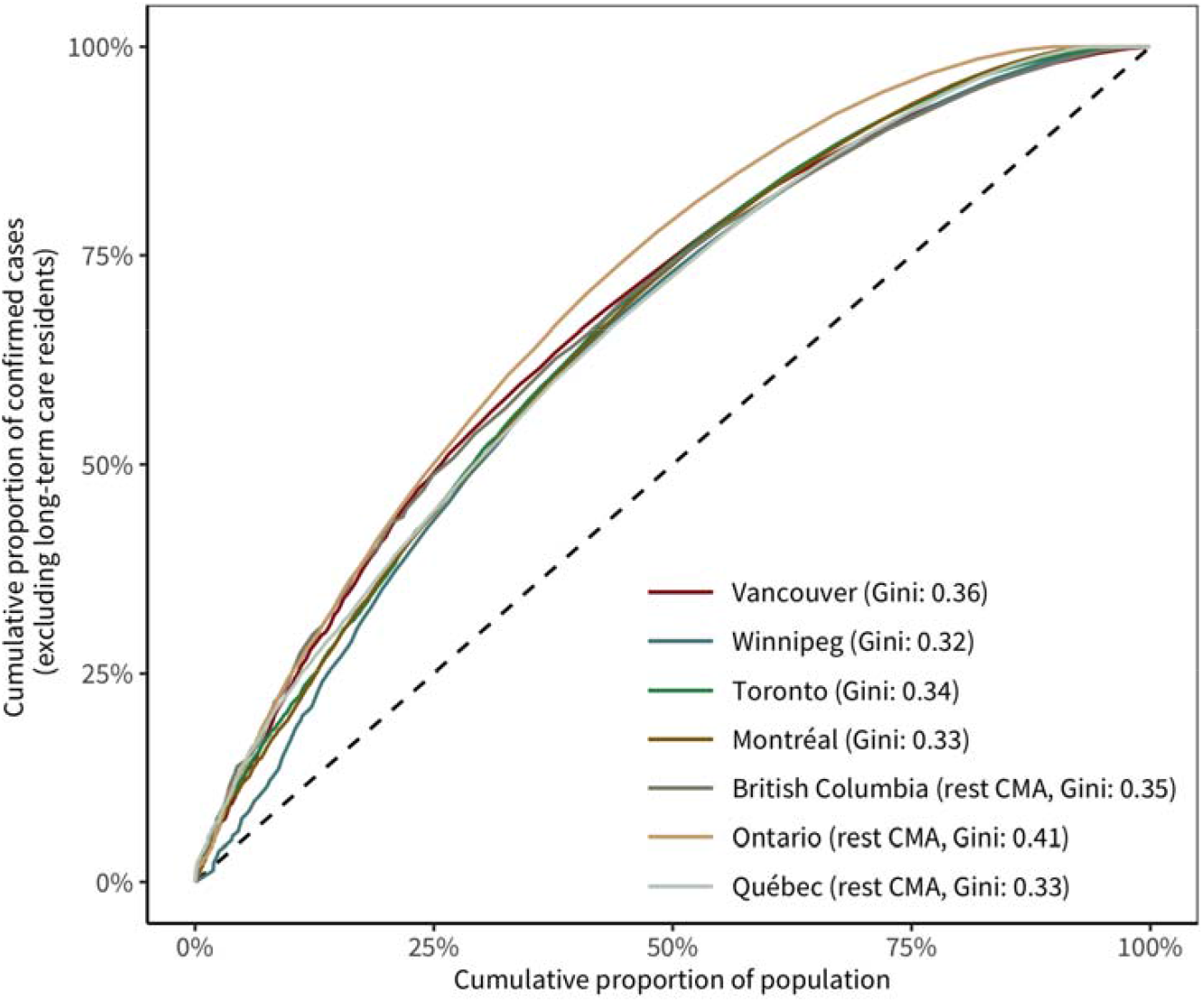
The Lorenz curves of COVID-19 confirmed cases (excluding long-term care residents) by proportion of the population and corresponding Gini coefficients. The population was ranked by the number of cases in each DA from the highest to the lowest. To ease interpretation, Abbotsford-Mission and Kelowna are grouped and displayed as “British Columbia (rest CMA)”; Kitchener-Cambridge– Waterloo, Hamilton, Ottawa-Gatineau (Ontario part), St. Catharines–Niagara and Windsor are grouped and displayed as “Ontario (rest CMA)”; Ottawa – Gatineau (Québec part), Québec City, Saguenay, Sherbrooke and Trois Rivières are grouped and displayed as “Québec (rest CMA)”. Lorenz curves and the corresponding Gini coefficients for each CMA can be found in *Figure S1*.

### Magnitude of heterogeneity by social determinants between cities

The social determinant across which nearly all CMAs experienced a concentration of cases was the proportion visible minority. **Figure 2** depicts the CMA-specific distribution and the respective co-Gini coefficients by proportion visible minority; concentration curves are depicted in **Figure 3**. Distribution of all the social determinants, co-Gini coefficients, proportion of population and the corresponding percentage of confirmed cases, and concentration curves for each CMA can be found in *Table S2, S3* and *Figure S2, S3*.

**Figure 2.**
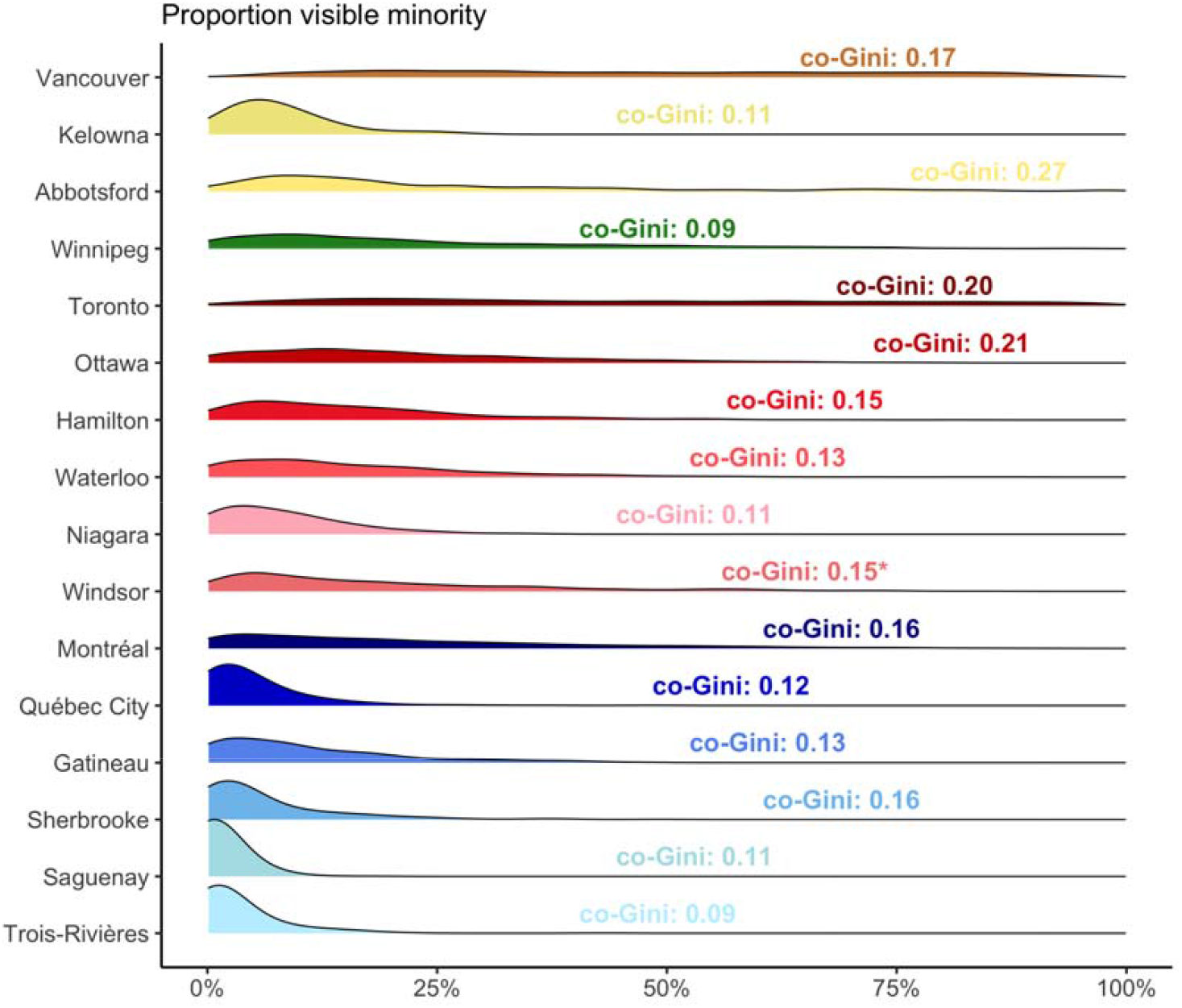
Distribution of proportion visible minority and the corresponding co-Gini coefficients (excluding long-term care residents) of cumulative COVID-19 cases across census metropolitan areas (CMA). Abbotsford-Mission is displayed as “Abbotsford”; Ottawa-Gatineau (Ontario part) is displayed as “Ottawa”; St. Catharines–Niagara is displayed as “Niagara”; Ottawa-Gatineau (Québec part) is displayed as “Gatineau”. Co-Gini coefficients followed by a “*” mark represent co-Gini coefficients of those Lorenz curves that went over and under the equality line. Distribution of other social determinants of health and the corresponding Gini (co-Gini) coefficients can be found in *Figure S2*.

**Figure 3.**
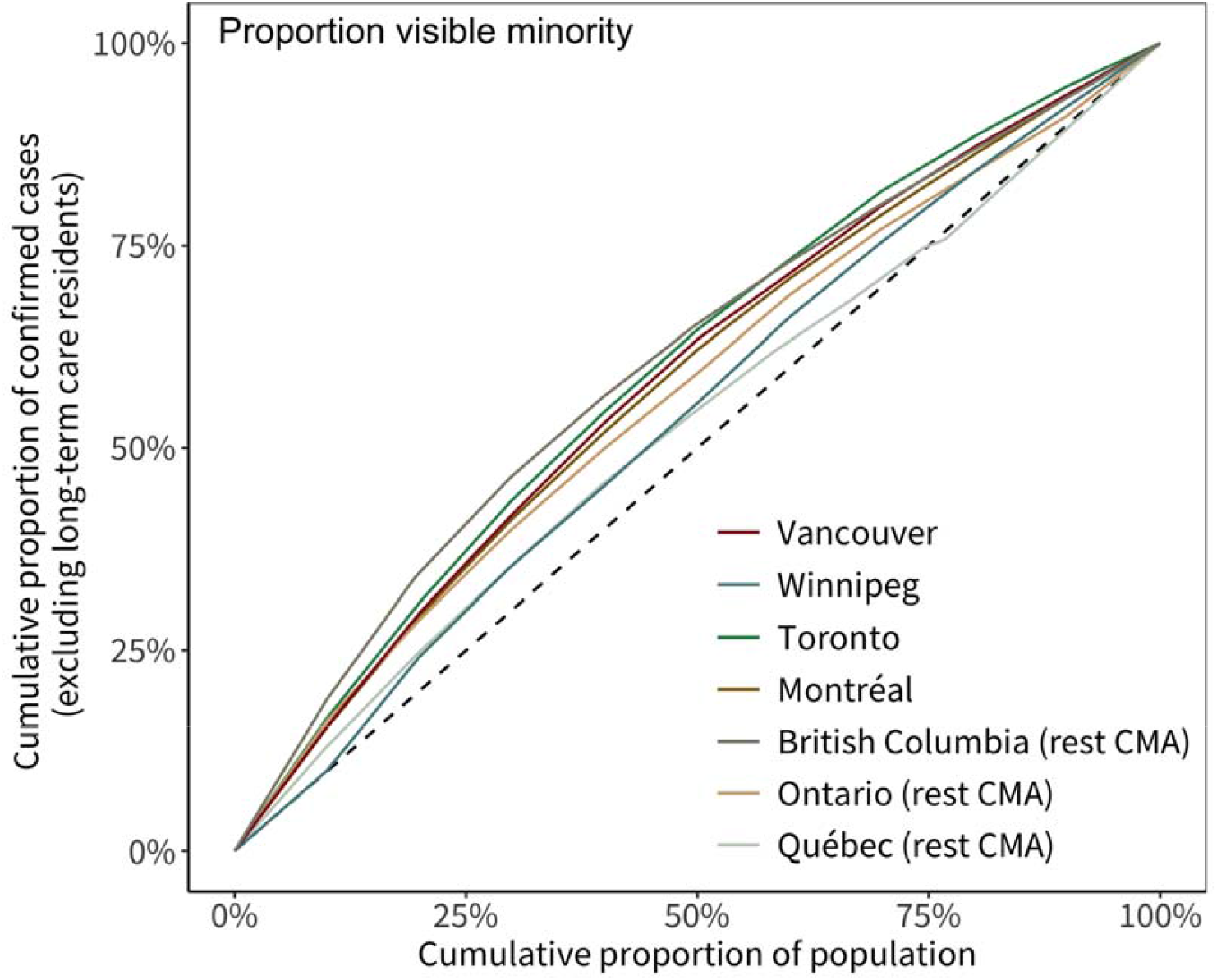
The concentration curves of COVID-19 confirmed cases (excluding long-term care residents) by proportion visible minority. The population was ranked by proportion visible minority from the highest decile to the lowest. To better visualize the figure, Abbotsford-Mission and Kelowna are grouped and displayed as “British Columbia (rest CMA)”; Kitchener-Cambridge–Waterloo, Hamilton, Ottawa-Gatineau (Ontario part), St. Catharines–Niagara and Windsor are grouped and displayed as “Ontario (rest CMA)”; Ottawa–Gatineau (Québec part), Québec City, Saguenay, Sherbrooke and Trois-Rivières are grouped and displayed as “Québec (rest CMA)”. Lorenz curves and Gini coefficients for each CMA can be found in *Figure S3*.

The distribution of the underlying social determinants was heterogenous across CMAs. Larger CMAs usually had wider distribution of the social determinants (**Figure S2**). Cities with less variability in the values of the social determinant tended to have smaller a co-Gini for that determinants: for example, Kelowna had a co-Gini of 0.07 for proportion suitable housing, whose distribution was narrow as compared with Vancouver (co-Gini 0.19, Figure S2). Across all CMAs, cases were disproportionately concentrated by geographies represented by lower income, smaller proportion of suitable housing, lower education attainment; and a higher proportion visible minority, recent immigration and essential workers (**Figure S3**). Concentration by visible minority was the most consistent finding across CMAs, with variability in inequalities across CMAs within provinces. The largest co-Gini coefficient for income was observed in Ottawa (co-Gini 0.17); for lower levels of education in Vancouver (0.24), for visible minority, recent immigration and suitable housing in Abbotsford-Mission (0.27, 0.23 and 0.21), and for essential workers in Vancouver (0.25) (**Table S2**). In Winnipeg (Manitoba), after-tax income explained the most heterogeneity (co-Gini 0.13).

When examining the 3 largest CMAs in Canada, the magnitude of geographical concentration by social determinants were similar for Toronto and Vancouver, in particular as they related to essential services (co-Gini 0.24 in Toronto, co-Gini 0.25 in Vancouver). In contrast, although Montréal demonstrated similar overall heterogeneity (Gini 0.33) to Toronto (0.34) and Vancouver (0.36), there was less heterogeneity by the same social determinants. In Montréal, the largest co-Gini was observed for proportion visible minority (co-Gini 0.16).

## Interpretation

This study provides comprehensive and robust evidence of high geographical concentration and thus, geographic hotspots of COVID-19 cases within Canadian cities across four provinces. These hotspots are largely defined along social determinants related to occupation, income, housing, and proxies for structural racism. Specifically, we quantified heterogeneities in cumulative COVID-19 cases using measures of inequality across sixteen Canadian CMAs from British Columbia, Manitoba, Ontario, and Québec – provinces with the majority of cases in Canada. Although the magnitude of geographical heterogeneity was relatively similar across CMAs, and a consistent theme across cities was the concentration of cases by proportion visibility minority, the degree of concentration by social determinants differed across cities.

There are two important implications of the findings for public health. First, given that each city demonstrated geographical concentration –with approximately 21-35% of the population accounting for 50% of cases– prioritizing and allocating resources to geographical hotspots could lead to a more effective and efficient response, and reduce inequalities (28), especially in the context of limited resources. An example of a hotspot-targeted strategy has been that of vaccination roll-out in some jurisdictions (29), but could also be systematically applied to ensure geographically-prioritized resources for timely access to testing, support for isolation and quarantine of contacts. Indeed, data suggest that without a systematic and intentional hotspot and community-tailored strategy, both testing and vaccination coverage were lowest in geographical hotspots (29, 30) and among visible minorities (24) in Canada and other high-income countries (31, 32). Second, given differences in the potential reasons for geographical heterogeneity between cities, each city would need to tailor its geographically-prioritized strategy to its local driver(s) of inequalities. For example, the difference in the co-Gini for essential services between Montréal compared to Vancouver and Toronto, despite similar distribution in the proportion essential workers in all three CMAs, suggests that the underlying context for hotspots (e.g., policies for sick leave (33)) may be different and thus signal different unmet needs of populations who shouldered the disproportionate burden of cases. Thus, using the spatial clustering of cases by social determinants to guide the local response could lead to more equitable allocation of resources and better access to interventions by providing services that actually meet the needs of communities at disproportionate risk. Such an approach may become even more important in the context of appropriately addressing the needs of ‘unvaccinated’ pockets of contact networks (34), and with increasingly transmissible variants of SARS-CoV-2 (35).

These results are consistent with the socio-geographical clustering patterns observed in other studies from Canada (24, 35), the United States (36-38), and Sweden (39). The social determinants examined in our study reflect those related to structural racism as measured via proportion visible minority and recent immigration (40, 41); occupational risks in the context of essential services with contacts rates, sometimes without occupational protections, and access to safe working environments (42, 43); limited educational attainment and its relationship with lower wages and barriers in access to accurate health information and healthcare (44, 45); and high-density households, especially as measured by lack of suitable housing which is a barrier to physical distancing and effective isolation or quarantine. Importantly, these determinants are often correlated (46). The concentration by social determinants reflects plausible mechanistic pathways for population-level transmission and, as such, the local contexts that define hotspots under broad stay-at-home policies (47) in each city.

Our descriptive study did not include an explanatory set of analyses to examine sources of heterogeneity in the difference in co-Gini between cities. However, we note that the distribution of each social determinant varied between CMAs, as depicted in **Figure S2**. When there is less variability of a given social determinant within a city, it consequently may be less of a determinant of geographical heterogeneity in cases. As such, the social determinants of geographical concentration between cities may also vary because of differences in the underlying level of homogeneity/heterogeneity by the determinant under study.

Limitations of our study include our use of observed cases, and thus we may underestimate the co-Gini if testing rates were lower among marginalized communities in particular, especially due to testing capacity constraints that were especially salient in the first wave (48-50). Second, although we excluded residents of long-term care homes, our definition of community-wide cases could still include other congregate-level settings such as shelters and group homes reflecting other ‘unmeasured’ determinants that could lead to geographical concentration within cities. Third, the DA-level social determinants were extracted from 2016 census data, which may not represent accurately the characteristics of the population in 2020-2021. Fourth, as individual-level data on social determinants for cases were not available, we conducted our unit of analysis at the smallest area (DA) possible to limit misclassification in the context of an ecological study. We limited the descriptive study to a cross-sectional analysis of each social determinant separately. Future work should examine sources of differences in the magnitude of inequalities/concentration in cases between cities (underlying differences in distribution of social determinants and the application of interventions), over time (to examine longitudinal pattern of heterogeneities over time and in each wave), and by a composite measure of social determinants or via multivariable analyses (given the potential for differential correlation between social determinants in each city).

In conclusion, geographical hotspots characterized by social determinants have been a consistent feature the COVID-19 pandemic across major urban centers in British Columbia, Manitoba, Ontario, and Québec. The pattern of epidemic concentration and thus, inequalities, by social determinants has varied between cities. Geographically-prioritized allocation of resources and services that are tailored to the local drivers of inequalities in acquisition and transmission risk offer a path forward in the public health response to SARS-CoV-2’s resurgence as vaccination programs are being scaled-up.

## Data Availability

COVID-19 confirmed case data used in this study is not publicly available. Data related to area-level characteristics is derived from 2016 Census Canada datasets, PCCF, and PCCF+ files.

## Authors’ contributions

YX, HM, GM, SB, SM, and MMG conceived of and designed the study. YX and HM developed the analysis plans, wrote the code, and coordinated code sharing across provincial teams. YX, HM, HVG, and MS conducted the statistical analysis. YX conducted the literature search, conducted the pooled analyses and generated the figures, and drafted the manuscript. GM drafted supplementary Table 1. NJ, DV, TW, AK, KY, RK, DLB, MB, SM, and MMG interpreted results, supported data curation, critically reviewed and edited the article.

## Acknowledgments

We acknowledge financial support from the *McGill Interdisciplinary Initiative in Infection and Immunity* (MI4; to MM-G), with seed funding from the *MUHC Foundation*, and a *Canadian Institutes of Health Research* (CIHR) grant (to SM, MM-G, NJ, AK, TW, SB). MM-G’s research program is supported by a *Canada Research Chair* (Tier 2) in *Population Health Modeling*. SM’s research program is supported by a *Canada Research Chair* (Tier 2) in *Mathematical Modeling and Program Science*. We thank Andrew Calzavera for outlining approach to generating the income variable; Dr. Sharon Straus and Dr. Jeff Kwong, and Dr. Maria Sunderam for helpful discussions. In Ontario, the reported COVID-19 cases were obtained from the provincial Case and Contact Management data as part of the Public Health Ontario Integrated Public Health Information System (iPHIS) via the Ontario COVID-19 Modelling Consensus Table and with approval from the University of Toronto Health Sciences Research Ethics Board (protocol no. 39253). The data were made available by the Ontario Ministry of Health and Long-Term Care (MOHLTC) to the Ontario Modelling Consensus Table on a daily basis. The analyses, conclusions, opinions and statements expressed herein are solely those of the authors and do not reflect those of the funding or data sources; no endorsement is intended or should be inferred. We acknowledge MCHP for use of data contained in the Manitoba Population Research Data Repository under project 2020-046 (HIPC No. 2020/2021-32). Data used in this study came from this repository which is housed at the MCHP, University of Manitoba and were derived from data provided by Manitoba Health, Seniors and Active Living, Winnipeg Regional Health Authority, and Statistics Canada. We acknowledge the assistance of the BC Centre for Disease Control, BC Ministry of Health and Regional Health Authority staff involved in data access, procurement, and management of case data in British Columbia.

All inferences, opinions, and conclusions drawn in this report are those of the authors, and do not reflect the opinions or policies of the Data Steward(s).

---

**Supplementary Table 1.**
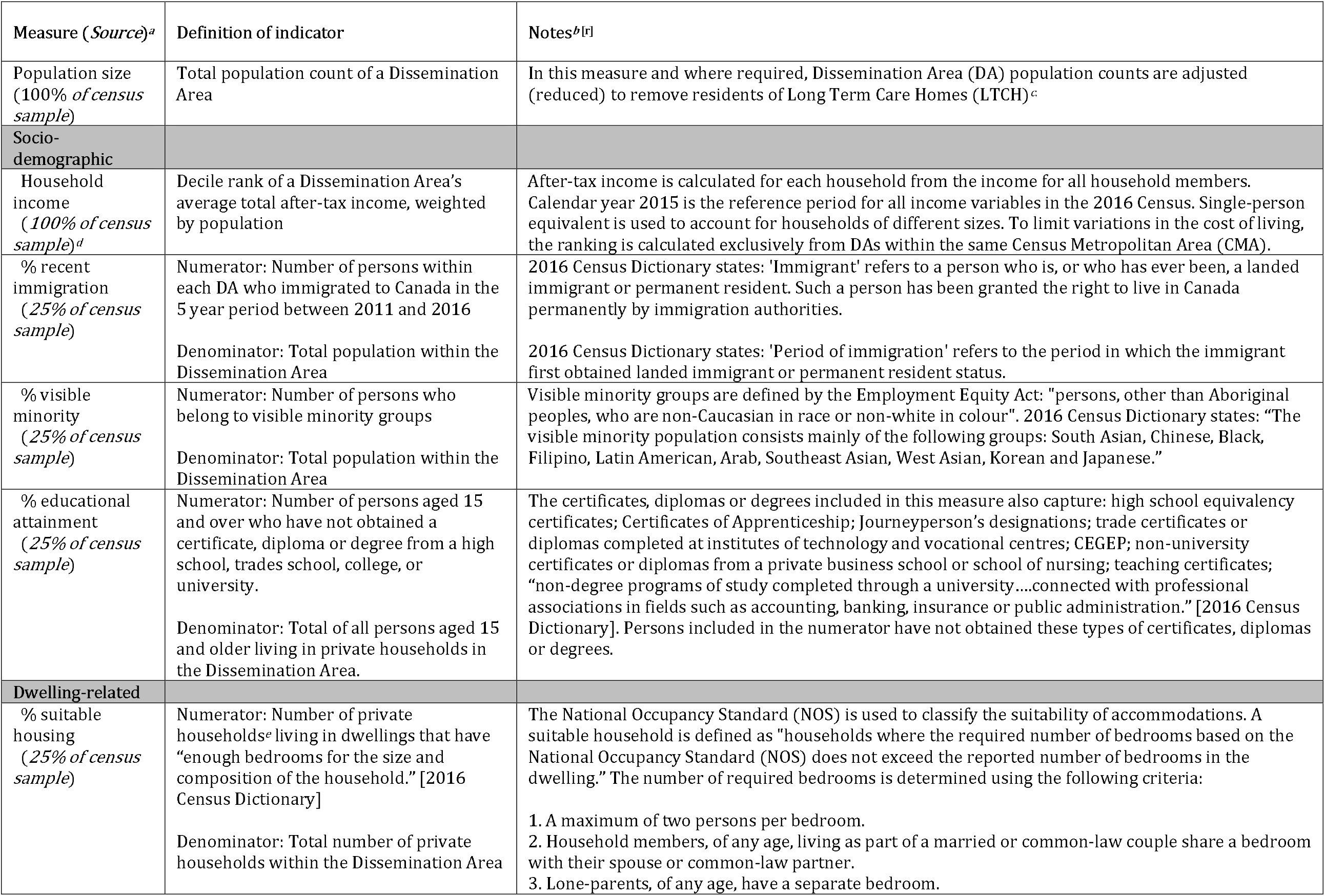
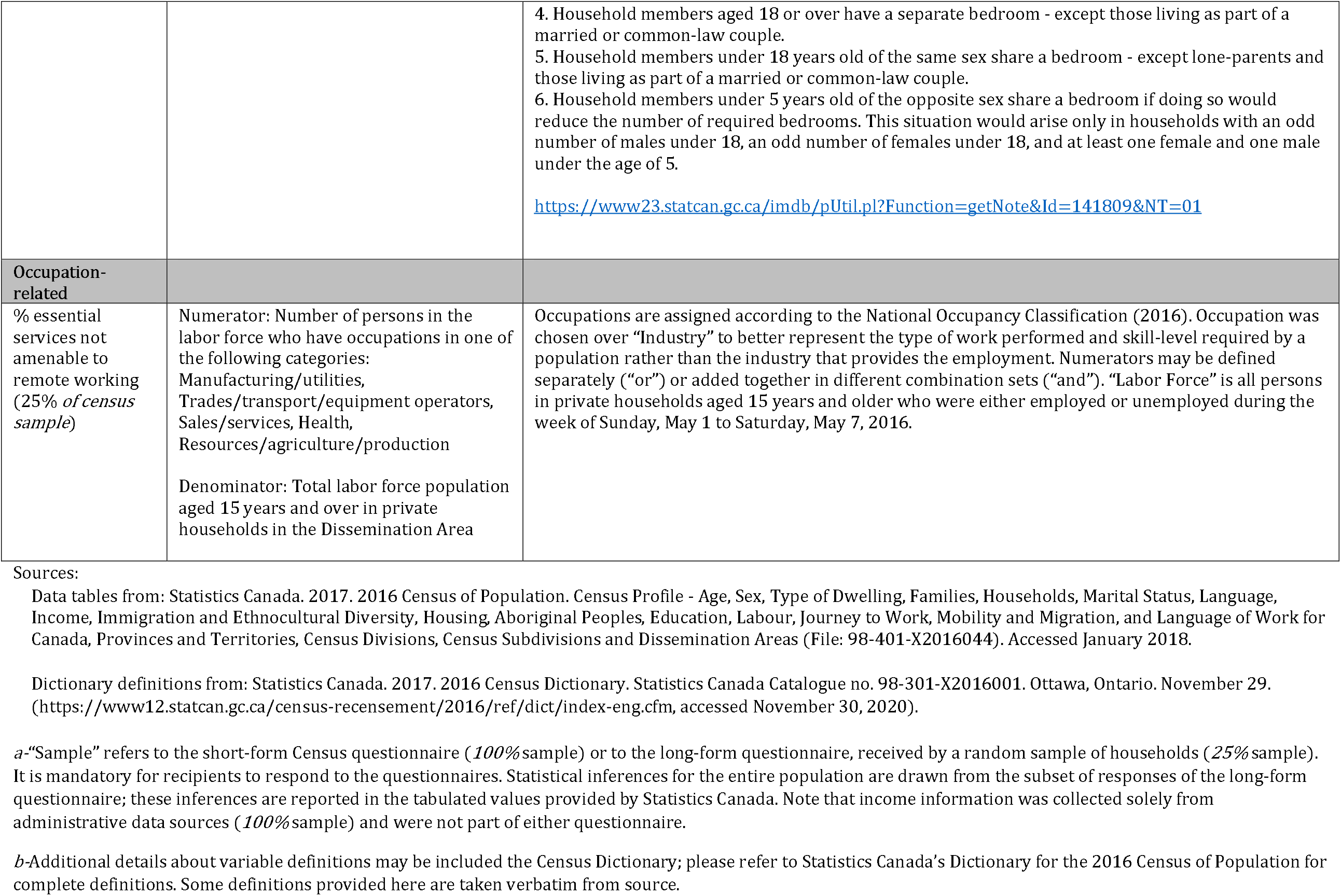
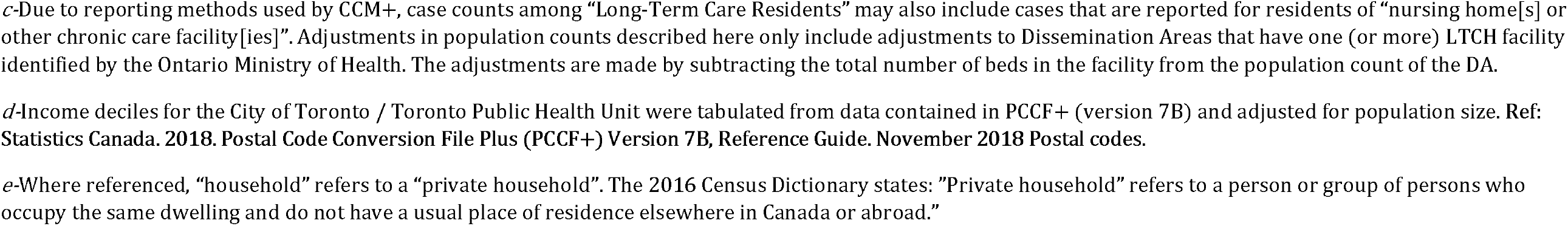
Social Determinants of Health – Variables from Statistics Canada 2016 Census of Population

**Table S2.**
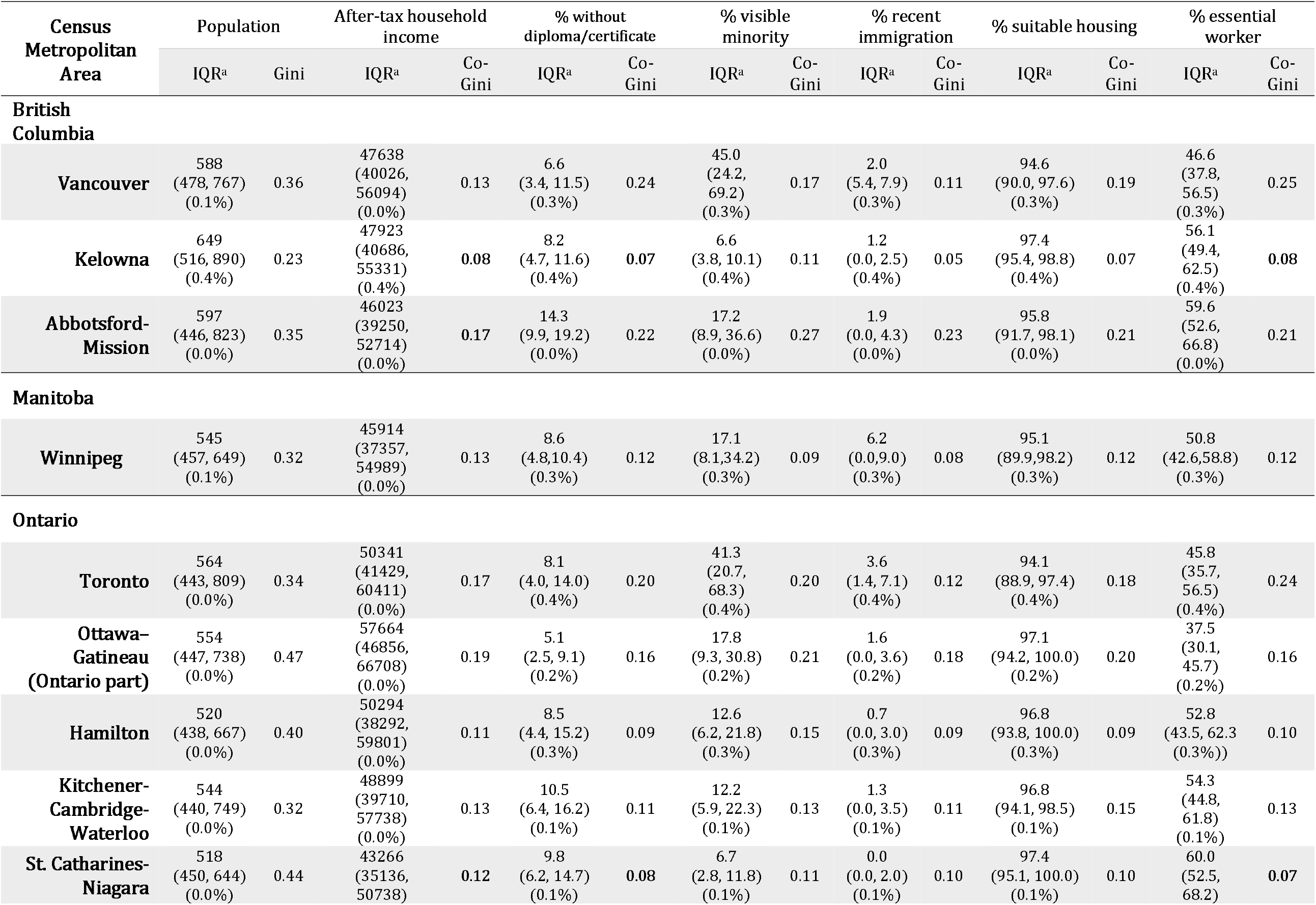
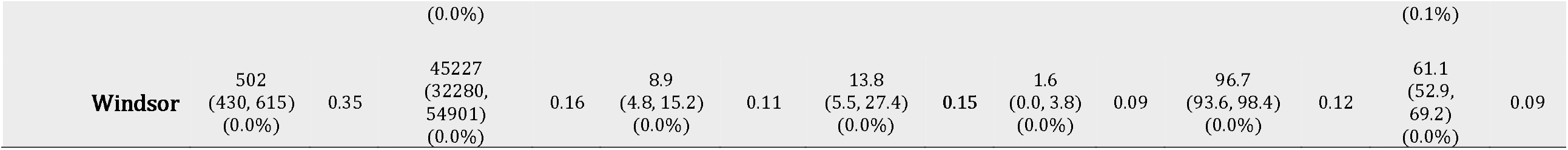
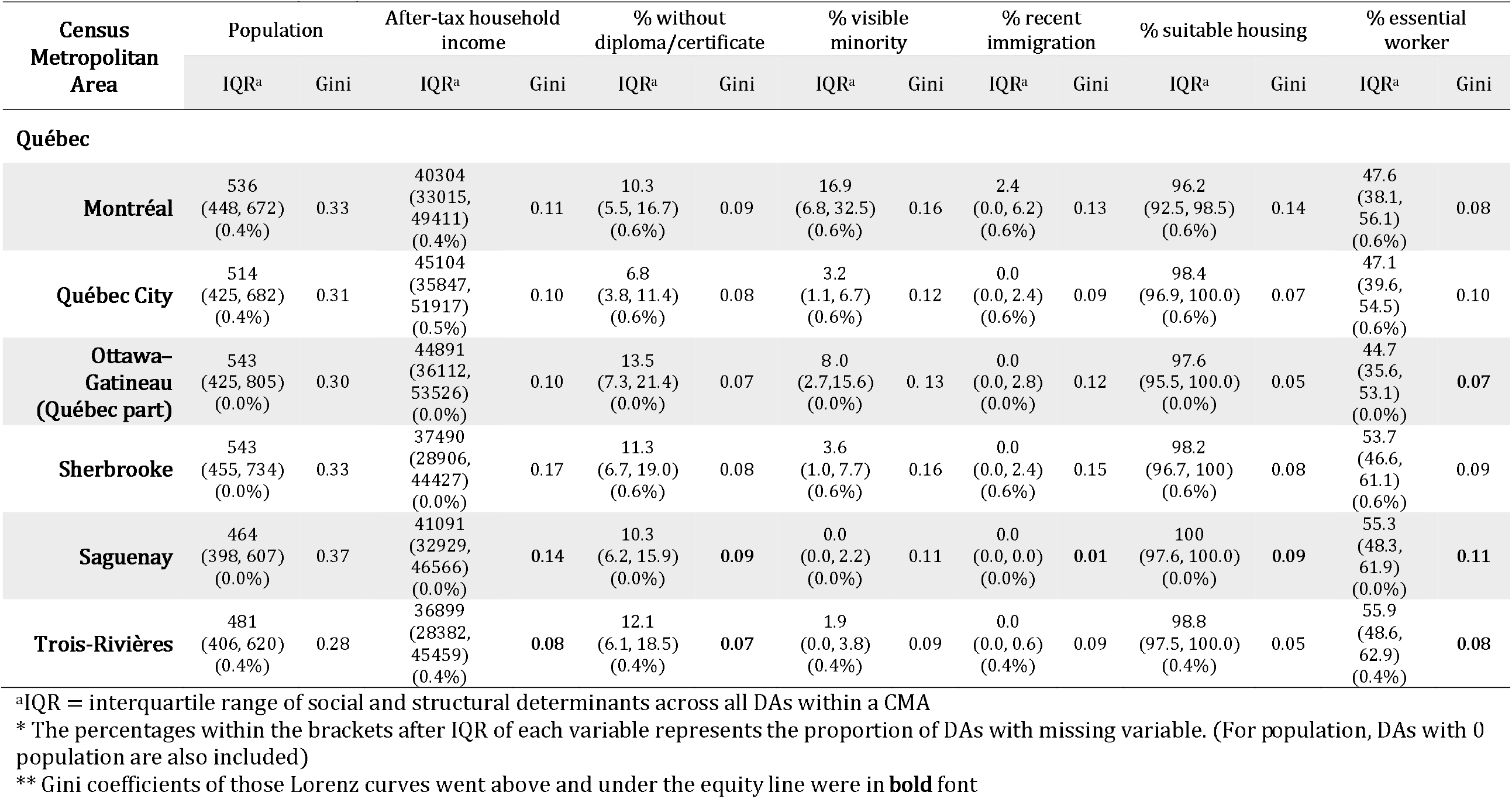
Characteristics of social and structural determinants across all dissemination area (DA) of each census metropolitan area (CMA) and the corresponding Gini/co-Gini coefficients of cumulative COVID-19 cases. All the variables are ranked from the highest value to the lowest.

**Table S3.** Proportion of population and the corresponding percentage of confirmed cases within each decile group ranked by the social and structural determinants across CMA.

| Census Metropolitan Area | Decile groups | After-tax household income |  | % without diploma/certificate |  | % visible minority |  | % recent immigration |  | % suitable housing |  | % essential worker |  |
| --- | --- | --- | --- | --- | --- | --- | --- | --- | --- | --- | --- | --- | --- |
|  |  | Pop* | Case** | Pop* | Case** | Pop* | Case** | Pop* | Case** | Pop* | Case** | Pop* | Case** |
| British Columbia |  |  |  |  |  |  |  |  |  |  |  |  |  |
| Vancouver | 1 | 10.0% | 6.1% | 9.9% | 21.3% | 10.0% | 15.5% | 10.0% | 13.4% | 1.8% | 1.0% | 10.0% | 23.7% |
|  | 2 | 20.0% | 13.0% | 20.0% | 37.3% | 20.0% | 29.6% | 20.0% | 25.7% | 19.7% | 13.0% | 20.0% | 38.8% |
|  | 3 | 30.0% | 20.9% | 30.0% | 49.5% | 30.0% | 41.8% | 29.9% | 37.0% | 29.9% | 21.1% | 30.0% | 49.3% |
|  | 4 | 40.0% | 29.9% | 40.0% | 58.7% | 39.9% | 53.1% | 40.0% | 47.6% | 39.9% | 29.0% | 39.3% | 58.3% |
|  | 5 | 50.0% | 39.6% | 49.9% | 66.6% | 50.0% | 63.4% | 50.0% | 57.9% | 49.3% | 37.1% | 50.0% | 66.5% |
|  | 6 | 60.0% | 51.0% | 60.0% | 73.8% | 60.0% | 71.6% | 60.0% | 68.6% | 59.8% | 47.0% | 59.9% | 73.6% |
|  | 7 | 70.0% | 63.3% | 69.6% | 80.5% | 70.0% | 80.0% | 70.0% | 77.1% | 69.9% | 57.3% | 70.0% | 80.4% |
|  | 8 | 80.0% | 76.4% | 79.7% | 87.0% | 80.0% | 87.3% | 79.9% | 85.4% | 79.9% | 68.1% | 80.0% | 87.2% |
|  | 9 | 90.0% | 89.0% | 89.9% | 94.2% | 90.0% | 93.8% | 88.6% | 92.0% | 89.9% | 81.1% | 89.8% | 93.8% |
|  | 10 | 100.0% | 100.0% | 100.0% | 100.0% | 100.0% | 100.0% | 100.0% | 100.0% | 100.0% | 100.0% | 100.0% | 100.0% |
| Kelowna | 1 | 9.8% | 10.5% | 9.9% | 12.3% | 9.9% | 14.4% | 9.4% | 11.0% | 2.7% | 2.2% | 9.6% | 10.7% |
|  | 2 | 19.8% | 22.0% | 19.9% | 21.6% | 19.1% | 25.1% | 19.6% | 21.8% | 19.2% | 16.8% | 19.9% | 20.1% |
|  | 3 | 29.8% | 32.5% | 29.1% | 30.8% | 30.0% | 37.4% | 30.0% | 31.7% | 29.7% | 26.9% | 29.8% | 30.5% |
|  | 4 | 39.8% | 41.5% | 40.0% | 39.7% | 39.7% | 45.7% | 39.1% | 40.5% | 39.1% | 37.0% | 39.9% | 39.8% |
|  | 5 | 49.8% | 51.4% | 49.5% | 48.6% | 49.5% | 55.7% | 49.2% | 49.4% | 49.4% | 46.7% | 49.8% | 47.6% |
|  | 6 | 59.6% | 58.6% | 59.8% | 58.9% | 59.8% | 65.8% | 59.7% | 58.9% | 59.9% | 58.0% | 59.7% | 58.6% |
|  | 7 | 69.9% | 67.7% | 69.2% | 66.6% | 69.5% | 74.1% | 64.9% | 64.7% | 69.2% | 66.4% | 68.0% | 66.6% |
|  | 8 | 79.3% | 76.3% | 79.7% | 78.0% | 79.0% | 83.6% | 100.0% | 100.0% | 79.9% | 76.8% | 79.7% | 80.7% |
|  | 9 | 89.8% | 88.6% | 89.7% | 88.9% | 89.9% | 92.4% |  |  | 89.8% | 88.5% | 89.9% | 88.9% |
|  | 10 | 100.0% | 100.0% | 100.0% | 100.0% | 100.0% | 100.0% |  |  | 100.0% | 100.0% | 100.0% | 100.0% |
| Abbotsford-Mission | 1 | 10.0% | 6.4% | 9.6% | 17.3% | 10.0% | 21.1% | 9.8% | 21.2% | 3.1% | 2.9% | 9.9% | 18.4% |
|  | 2 | 19.9% | 12.0% | 19.7% | 35.1% | 19.9% | 38.6% | 19.7% | 34.5% | 19.3% | 13.5% | 19.9% | 32.5% |
|  | 3 | 30.0% | 20.3% | 30.0% | 46.6% | 29.5% | 50.9% | 29.8% | 44.8% | 29.5% | 20.1% | 29.7% | 46.2% |
|  | 4 | 39.7% | 28.9% | 39.8% | 56.0% | 40.0% | 61.7% | 40.0% | 56.2% | 39.6% | 26.9% | 39.9% | 56.8% |
|  | 5 | 49.9% | 38.6% | 49.6% | 65.4% | 49.8% | 69.9% | 49.5% | 64.0% | 49.1% | 34.8% | 49.4% | 64.4% |
|  | 6 | 59.7% | 51.4% | 59.7% | 74.2% | 59.4% | 76.3% | 59.8% | 73.4% | 59.9% | 43.2% | 59.6% | 72.5% |
|  | 7 | 69.7% | 66.6% | 70.0% | 80.8% | 70.0% | 83.0% | 69.4% | 81.1% | 68.4% | 54.7% | 69.9% | 80.9% |
|  | 8 | 79.8% | 81.5% | 79.9% | 88.1% | 79.8% | 88.1% | 73.6% | 83.3% | 79.7% | 66.3% | 79.9% | 87.4% |
|  | 9 | 89.8% | 90.8% | 89.3% | 94.1% | 89.9% | 93.8% | 100.0% | 100.0% | 89.9% | 79.8% | 89.9% | 93.8% |
|  | 10 | 100.0% | 100.0% | 100.0% | 100.0% | 100.0% | 100.0% |  |  | 100.0% | 100.0% | 100.0% | 100.0% |

**Table S3 (Cont.)**
| Census Metropolitan Area | Decile groups | After-tax household income |  | % without diploma/certificate |  | % visible minority |  | % recent immigration |  | % suitable housing |  | % essential worker |  |
| --- | --- | --- | --- | --- | --- | --- | --- | --- | --- | --- | --- | --- | --- |
|  |  | Pop* | Case** | Pop* | Case** | Pop* | Case** | Pop* | Case** | Pop* | Case** | Pop* | Case** |
| Manitoba |  |  |  |  |  |  |  |  |  |  |  |  |  |
| Winnipeg | 1 | 9.9% | 7.4% | 10.0% | 15.6% | 9.9% | 9.8% | 9.9% | 12.4% | 2.4% | 2.1% | 10.0% | 14.3% |
|  | 2 | 19.8% | 14.4% | 20.0% | 28.1% | 19.9% | 23.8% | 19.9% | 23.4% | 19.9% | 16.2% | 19.7% | 27.0% |
|  | 3 | 30.0% | 22.8% | 29.9% | 38.6% | 29.7% | 35.0% | 30.0% | 35.0% | 29.6% | 24.0% | 30.0% | 39.0% |
|  | 4 | 40.0% | 29.9% | 39.9% | 48.5% | 39.9% | 45.2% | 39.9% | 45.7% | 39.9% | 31.6% | 40.0% | 48.9% |
|  | 5 | 50.0% | 39.2% | 49.9% | 57.9% | 50.0% | 55.8% | 49.9% | 56.4% | 49.9% | 40.9% | 49.1% | 57.8% |
|  | 6 | 59.9% | 49.7% | 59.8% | 67.0% | 59.9% | 66.8% | 59.6% | 65.1% | 60.0% | 51.1% | 60.0% | 67.8% |
|  | 7 | 70.0% | 60.6% | 69.6% | 75.7% | 69.9% | 76.4% | 69.9% | 75.2% | 69.5% | 59.8% | 70.0% | 75.1% |
|  | 8 | 79.7% | 72.8% | 80.0% | 82.7% | 80.0% | 85.5% | 77.9% | 80.9% | 79.9% | 72.0% | 80.0% | 83.4% |
|  | 9 | 89.8% | 84.6% | 90.0% | 90.9% | 90.0% | 93.5% | 100.0% | 100.0% | 89.6% | 85.2% | 89.9% | 91.0% |
|  | 10 | 100.0% | 100.0% | 100.0% | 100.0% | 100.0% | 100.0% |  |  | 100.0% | 100.0% | 100.0% | 100.0% |

Table S3 (Cont.)
| Census Metropolitan Area | Decile groups | After-tax household income |  | % without diploma/certificate |  | % visible minority |  | % recent immigration |  | % suitable housing |  | % essential worker |  |
| --- | --- | --- | --- | --- | --- | --- | --- | --- | --- | --- | --- | --- | --- |
|  |  | Pop* | Case** | Pop* | Case** | Pop* | Case** | Pop* | Case** | Pop* | Case** | Pop* | Case** |
| Ontario |  |  |  |  |  |  |  |  |  |  |  |  |  |
| Toronto | 1 | 10.0% | 4.9% | 9.8% | 15.4% | 10.0% | 16.6% | 9.9% | 13.4% | 1.6% | 1.0% | 10.0% | 18.1% |
|  | 2 | 20.0% | 11.0% | 19.9% | 30.2% | 20.0% | 30.9% | 20.0% | 25.6% | 19.7% | 11.7% | 20.0% | 33.1% |
|  | 3 | 30.0% | 18.5% | 29.9% | 43.0% | 30.0% | 43.6% | 29.8% | 37.7% | 29.9% | 19.0% | 29.9% | 46.7% |
|  | 4 | 40.0% | 27.2% | 39.7% | 54.6% | 40.0% | 54.5% | 40.0% | 48.4% | 39.9% | 26.8% | 39.9% | 58.4% |
|  | 5 | 50.0% | 37.4% | 50.0% | 65.5% | 50.0% | 64.6% | 49.8% | 58.9% | 50.0% | 36.2% | 50.0% | 67.9% |
|  | 6 | 60.0% | 49.4% | 59.9% | 74.6% | 59.9% | 73.2% | 60.0% | 69.2% | 59.9% | 46.3% | 60.0% | 76.7% |
|  | 7 | 70.0% | 61.2% | 69.9% | 82.1% | 70.0% | 81.8% | 69.9% | 78.3% | 70.0% | 57.5% | 70.0% | 84.2% |
|  | 8 | 80.0% | 72.8% | 79.8% | 88.4% | 80.0% | 88.6% | 80.0% | 86.4% | 79.9% | 69.9% | 79.9% | 90.1% |
|  | 9 | 90.0% | 85.1% | 89.9% | 95.0% | 90.0% | 94.8% | 85.8% | 90.5% | 90.0% | 83.9% | 90.0% | 95.4% |
|  | 10 | 100.0% | 100.0% | 100.0% | 100.0% | 100.0% | 100.0% | 100.0% | 100.0% | 100.0% | 100.0% | 100.0% | 100.0% |
| Ottawa – Gatineau (Ontario part) | 1 | 9.9% | 5.7% | 10.0% | 17.6% | 10.0% | 20.2% | 9.9% | 19.2% | 2.9% | 1.8% | 10.0% | 19.1% |
|  | 2 | 19.9% | 12.1% | 20.0% | 31.3% | 20.0% | 33.0% | 20.0% | 30.7% | 29.6% | 19.2% | 20.0% | 30.5% |
|  | 3 | 30.0% | 19.3% | 29.9% | 42.2% | 29.9% | 44.7% | 29.8% | 43.1% | 39.6% | 27.8% | 30.0% | 39.7% |
|  | 4 | 40.0% | 27.7% | 39.4% | 50.3% | 40.0% | 55.0% | 40.0% | 53.0% | 49.6% | 36.1% | 39.7% | 50.3% |
|  | 5 | 50.0% | 37.8% | 49.8% | 61.6% | 50.0% | 65.0% | 49.9% | 63.9% | 59.9% | 45.4% | 50.0% | 60.6% |
|  | 6 | 59.9% | 48.0% | 59.9% | 70.0% | 59.8% | 73.5% | 60.0% | 71.3% | 70.0% | 55.7% | 60.0% | 69.4% |
|  | 7 | 70.0% | 56.7% | 68.7% | 76.7% | 69.9% | 81.8% | 66.2% | 76.1% | 80.0% | 67.4% | 70.0% | 78.1% |
|  | 8 | 79.9% | 67.7% | 80.0% | 85.8% | 80.0% | 88.5% | 100.0% | 100.0% | 89.9% | 79.2% | 79.9% | 86.8% |
|  | 9 | 90.0% | 80.9% | 84.1% | 89.1% | 90.0% | 94.0% |  |  | 100.0% | 100.0% | 90.0% | 94.0% |
|  | 10 | 100.0% | 100.0% | 100.0% | 100.0% | 100.0% | 100.0% |  |  |  |  | 100.0% | 100.0% |
| Hamilton | 1 | 9.9% | 6.9% | 10.0% | 11.5% | 10.0% | 16.3% | 9.9% | 12.5% | 2.5% | 2.1% | 9.9% | 13.0% |
|  | 2 | 20.0% | 15.1% | 19.8% | 23.3% | 19.9% | 28.7% | 20.0% | 24.3% | 29.6% | 25.3% | 20.0% | 22.7% |
|  | 3 | 29.9% | 23.9% | 30.0% | 35.0% | 30.0% | 39.2% | 29.9% | 34.9% | 39.4% | 33.3% | 30.0% | 34.2% |
|  | 4 | 40.0% | 31.7% | 39.9% | 44.1% | 40.0% | 48.9% | 39.8% | 46.7% | 49.4% | 43.0% | 39.9% | 45.0% |
|  | 5 | 49.9% | 44.4% | 49.7% | 56.5% | 49.9% | 56.8% | 49.9% | 56.1% | 59.4% | 53.8% | 50.0% | 57.0% |
|  | 6 | 60.0% | 54.4% | 59.8% | 65.7% | 60.0% | 68.0% | 58.7% | 63.2% | 69.3% | 65.0% | 59.9% | 65.5% |
|  | 7 | 70.0% | 65.3% | 69.8% | 74.5% | 69.9% | 78.0% | 100.0% | 100.0% | 79.9% | 75.7% | 69.9% | 75.5% |
|  | 8 | 80.0% | 75.8% | 79.8% | 83.2% | 80.0% | 85.2% |  |  | 89.5% | 85.3% | 80.0% | 84.1% |
|  | 9 | 89.9% | 87.9% | 89.9% | 91.2% | 90.0% | 92.0% |  |  | 100.0% | 100.0% | 90.0% | 91.4% |
|  | 10 | 100.0% | 100.0% | 100.0% | 100.0% | 100.0% | 100.0% |  |  |  |  | 100.0% | 100.0% |

Table S3 (Cont.)
| Census Metropolitan Area | Decile groups | After-tax household income |  | % without diploma/certificate |  | % visible minority |  | % recent immigration |  | % suitable housing |  | % essential worker |  |
| --- | --- | --- | --- | --- | --- | --- | --- | --- | --- | --- | --- | --- | --- |
|  |  | Pop* | Case** | Pop* | Case** | Pop* | Case** | Pop* | Case** | Pop* | Case** | Pop* | Case** |
| Kitchener - Cambridge - Waterloo | 1 | 10.0% | 7.4% | 9.9% | 15.2% | 10.0% | 15.2% | 9.9% | 12.6% | 3.1% | 1.8% | 9.9% | 14.7% |
|  | 2 | 20.0% | 15.8% | 19.7% | 25.7% | 19.6% | 26.5% | 19.9% | 23.8% | 20.0% | 15.4% | 19.9% | 26.8% |
|  | 3 | 30.0% | 22.8% | 29.7% | 37.4% | 30.0% | 37.8% | 29.8% | 36.6% | 29.9% | 23.5% | 29.9% | 38.0% |
|  | 4 | 40.0% | 31.5% | 39.9% | 48.6% | 40.0% | 47.2% | 39.8% | 46.8% | 39.8% | 32.4% | 40.0% | 48.5% |
|  | 5 | 49.2% | 41.0% | 49.9% | 57.8% | 50.0% | 57.5% | 49.8% | 56.8% | 49.9% | 42.8% | 49.5% | 57.8% |
|  | 6 | 59.9% | 51.0% | 59.8% | 66.3% | 59.9% | 66.7% | 59.8% | 65.8% | 59.4% | 50.6% | 59.9% | 67.8% |
|  | 7 | 70.0% | 61.6% | 69.9% | 75.4% | 70.0% | 75.1% | 65.0% | 69.2% | 69.9% | 62.1% | 70.0% | 77.8% |
|  | 8 | 79.9% | 71.4% | 79.4% | 83.7% | 80.0% | 81.7% | 100.0% | 100.0% | 80.0% | 72.4% | 79.8% | 84.2% |
|  | 9 | 89.9% | 83.6% | 89.8% | 92.7% | 90.0% | 89.1% |  |  | 89.8% | 82.1% | 90.0% | 92.5% |
|  | 10 | 100.0% | 100.0% | 100.0% | 100.0% | 100.0% | 100.0% |  |  | 100.0% | 100.0% | 100.0% | 100.0% |
| St. Catharines - Niagara | 1 | 10.0% | 14.3% | 10.0% | 9.2% | 9.7% | 12.5% | 9.9% | 12.0% | 4.2% | 3.0% | 9.9% | 10.9% |
|  | 2 | 19.9% | 21.9% | 19.8% | 20.5% | 19.8% | 25.6% | 19.6% | 23.8% | 29.9% | 28.3% | 20.0% | 21.9% |
|  | 3 | 29.8% | 29.1% | 29.5% | 30.9% | 30.0% | 36.9% | 30.0% | 36.7% | 39.3% | 36.3% | 29.9% | 32.2% |
|  | 4 | 39.9% | 37.4% | 39.9% | 41.1% | 39.7% | 46.6% | 39.7% | 47.7% | 49.5% | 43.9% | 40.0% | 40.1% |
|  | 5 | 50.0% | 49.5% | 49.9% | 50.2% | 49.9% | 56.4% | 42.8% | 49.9% | 59.8% | 55.3% | 49.9% | 50.9% |
|  | 6 | 59.9% | 59.4% | 59.8% | 57.8% | 60.0% | 66.7% | 100.0% | 100.0% | 69.7% | 66.3% | 59.9% | 60.6% |
|  | 7 | 70.0% | 69.1% | 70.0% | 68.6% | 69.9% | 73.2% |  |  | 79.5% | 76.0% | 69.9% | 69.8% |
|  | 8 | 79.9% | 77.9% | 79.7% | 77.1% | 79.7% | 81.3% |  |  | 89.9% | 87.0% | 79.9% | 77.8% |
|  | 9 | 89.9% | 88.1% | 89.9% | 90.8% | 88.8% | 91.2% |  |  | 100.0% | 100.0% | 89.7% | 87.3% |
|  | 10 | 100.0% | 100.0% | 100.0% | 100.0% | 100.0% | 100.0% |  |  |  |  | 100.0% | 100.0% |
| Windsor | 1 | 9.8% | 6.8% | 10.0% | 13.4% | 10.0% | 13.8% | 9.9% | 13.4% | 2.1% | 1.4% | 10.0% | 11.1% |
|  | 2 | 19.8% | 14.5% | 19.3% | 25.7% | 19.9% | 26.3% | 19.8% | 25.3% | 29.8% | 25.9% | 19.9% | 23.7% |
|  | 3 | 29.7% | 21.6% | 29.8% | 36.2% | 29.9% | 38.1% | 29.8% | 35.6% | 38.9% | 33.6% | 30.0% | 33.5% |
|  | 4 | 39.7% | 34.0% | 39.7% | 46.4% | 39.9% | 48.8% | 39.9% | 45.9% | 49.8% | 42.5% | 39.4% | 43.1% |
|  | 5 | 49.7% | 44.9% | 50.0% | 55.2% | 49.6% | 57.2% | 50.0% | 54.4% | 59.8% | 53.0% | 49.7% | 51.5% |
|  | 6 | 59.9% | 57.8% | 59.9% | 63.6% | 60.0% | 66.5% | 59.5% | 64.8% | 69.9% | 62.3% | 60.0% | 63.9% |
|  | 7 | 69.9% | 64.9% | 69.8% | 72.1% | 69.6% | 73.6% | 100.0% | 100.0% | 79.9% | 73.1% | 69.9% | 72.4% |
|  | 8 | 80.0% | 73.8% | 79.8% | 79.8% | 79.8% | 80.2% |  |  | 89.9% | 86.6% | 79.9% | 83.4% |
|  | 9 | 89.9% | 84.0% | 89.8% | 91.8% | 89.6% | 86.4% |  |  | 100.0% | 100.0% | 89.9% | 91.0% |
|  | 10 | 100.0% | 100.0% | 100.0% | 100.0% | 100.0% | 100.0% |  |  |  |  | 100.0% | 100.0% |

Table S3 (Cont.)
| Census Metropolitan Area | Decile groups | After-tax household income |  | % without diploma/certificate |  | % visible minority |  | % recent immigration |  | % suitable housing |  | % essential worker |  |
| --- | --- | --- | --- | --- | --- | --- | --- | --- | --- | --- | --- | --- | --- |
|  |  | Pop* | Case** | Pop* | Case** | Pop* | Case** | Pop* | Case** | Pop* | Case** | Pop* | Case** |
| Québec |  |  |  |  |  |  |  |  |  |  |  |  |  |
| Montreal | 1 | 9.9% | 6.8% | 10.0% | 12.8% | 10.0% | 15.5% | 10.0% | 14.9% | 2.6% | 2.0% | 10.0% | 11.8% |
|  | 2 | 20.0% | 15.3% | 20.0% | 24.5% | 20.0% | 29.2% | 20.0% | 26.6% | 20.0% | 15.1% | 20.0% | 23.4% |
|  | 3 | 30.0% | 23.9% | 29.7% | 35.0% | 30.0% | 41.2% | 29.9% | 37.7% | 29.9% | 22.8% | 29.8% | 34.5% |
|  | 4 | 40.0% | 32.5% | 40.0% | 45.8% | 40.0% | 52.0% | 40.0% | 47.9% | 39.5% | 31.1% | 38.5% | 43.7% |
|  | 5 | 50.0% | 42.0% | 49.8% | 55.8% | 50.0% | 62.1% | 50.0% | 57.9% | 49.6% | 39.5% | 50.0% | 55.7% |
|  | 6 | 60.0% | 51.8% | 60.0% | 66.0% | 60.0% | 70.9% | 60.0% | 67.0% | 60.0% | 49.2% | 59.9% | 65.5% |
|  | 7 | 70.0% | 62.4% | 70.0% | 75.1% | 69.9% | 78.9% | 70.0% | 75.6% | 69.9% | 59.1% | 70.0% | 75.5% |
|  | 8 | 80.0% | 74.4% | 79.9% | 84.3% | 80.0% | 86.4% | 70.6% | 75.9% | 80.0% | 70.9% | 79.9% | 83.9% |
|  | 9 | 90.0% | 86.7% | 90.0% | 92.5% | 90.0% | 93.4% | 100.0% | 100.0% | 89.7% | 84.0% | 89.9% | 92.2% |
|  | 10 | 100.0% | 100.0% | 100.0% | 100.0% | 100.0% | 100.0% |  |  | 100.0% | 100.0% | 100.0% | 100.0% |
| Quebec City | 1 | 10.0% | 7.6% | 10.0% | 13.5% | 9.9% | 13.1% | 9.9% | 12.2% | 6.3% | 6.0% | 10.0% | 12.3% |
|  | 2 | 19.9% | 15.8% | 19.9% | 24.4% | 20.0% | 24.3% | 20.0% | 24.3% | 39.7% | 36.6% | 19.9% | 22.7% |
|  | 3 | 30.0% | 24.7% | 29.5% | 34.4% | 30.0% | 35.2% | 29.6% | 34.0% | 49.9% | 45.0% | 30.0% | 35.7% |
|  | 4 | 40.0% | 33.7% | 39.9% | 44.5% | 40.0% | 45.3% | 39.9% | 43.6% | 59.9% | 55.3% | 40.0% | 45.7% |
|  | 5 | 50.0% | 42.9% | 49.8% | 54.5% | 49.8% | 54.2% | 47.2% | 49.1% | 70.0% | 66.7% | 49.9% | 54.6% |
|  | 6 | 60.0% | 52.3% | 59.6% | 64.2% | 59.9% | 63.1% | 100.0% | 100.0% | 80.0% | 76.0% | 59.9% | 66.4% |
|  | 7 | 70.0% | 62.1% | 70.0% | 74.5% | 70.0% | 70.9% |  |  | 89.7% | 87.1% | 69.9% | 75.0% |
|  | 8 | 80.0% | 73.7% | 79.7% | 82.8% | 79.9% | 79.5% |  |  | 100.0% | 100.0% | 79.9% | 83.8% |
|  | 9 | 90.0% | 86.5% | 88.3% | 89.5% | 80.5% | 79.8% |  |  |  |  | 90.0% | 92.2% |
|  | 10 | 100.0% | 100.0% | 100.0% | 100.0% | 100.0% | 100.0% |  |  |  |  | 100.0% | 100.0% |
| Ottawa – Gatineau (Quebec part) | 1 | 9.9% | 7.9% | 10.0% | 13.0% | 9.9% | 13.8% | 10.0% | 16.2% | 2.4% | 2.7% | 9.9% | 9.3% |
|  | 2 | 19.9% | 15.3% | 19.9% | 22.1% | 20.0% | 27.1% | 19.8% | 26.5% | 29.7% | 29.5% | 19.9% | 18.3% |
|  | 3 | 29.6% | 25.1% | 29.8% | 30.8% | 30.0% | 35.7% | 29.8% | 36.7% | 39.5% | 37.7% | 27.9% | 26.9% |
|  | 4 | 39.7% | 34.5% | 40.0% | 40.8% | 40.0% | 47.4% | 39.8% | 46.1% | 49.4% | 46.8% | 39.9% | 37.8% |
|  | 5 | 49.9% | 44.7% | 49.8% | 51.0% | 50.0% | 56.5% | 49.7% | 57.3% | 59.5% | 57.5% | 49.8% | 46.6% |
|  | 6 | 59.8% | 53.7% | 59.8% | 61.7% | 59.9% | 67.1% | 54.2% | 61.0% | 69.6% | 66.6% | 60.0% | 60.4% |
|  | 7 | 69.9% | 64.8% | 69.8% | 72.0% | 69.9% | 75.8% | 100.0% | 100.0% | 79.8% | 77.9% | 69.7% | 70.8% |
|  | 8 | 80.0% | 73.7% | 79.8% | 82.6% | 79.9% | 85.6% |  |  | 90.0% | 88.7% | 79.7% | 81.3% |
|  | 9 | 90.0% | 87.1% | 90.0% | 91.9% | 89.8% | 92.0% |  |  | 100.0% | 100.0% | 89.9% | 91.2% |
|  | 10 | 100.0% | 100.0% | 100.0% | 100.0% | 100.0% | 100.0% |  |  |  |  | 100.0% | 100.0% |

Table S3 (Cont.)
| Census Metropolitan Area | Decile groups | After-tax household income |  | % without diploma/certificate |  | % visible minority |  | % recent immigration |  | % suitable housing |  | % essential worker |  |
| --- | --- | --- | --- | --- | --- | --- | --- | --- | --- | --- | --- | --- | --- |
|  |  | Pop* | Case** | Pop* | Case** | Pop* | Case** | Pop* | Case** | Pop* | Case** | Pop* | Case** |
| Sherbrooke | 1 | 9.8% | 5.1% | 9.9% | 12.4% | 9.7% | 14.7% | 9.9% | 14.4% | 3.1% | 3.5% | 9.3% | 9.3% |
|  | 2 | 19.2% | 16.3% | 19.9% | 23.7% | 19.7% | 29.5% | 19.9% | 29.3% | 39.8% | 35.9% | 19.8% | 22.7% |
|  | 3 | 29.9% | 24.9% | 29.9% | 35.2% | 29.6% | 38.4% | 29.6% | 36.5% | 50.0% | 44.4% | 29.6% | 31.6% |
|  | 4 | 39.9% | 31.5% | 39.9% | 46.0% | 38.9% | 48.1% | 39.6% | 47.1% | 59.2% | 53.5% | 39.6% | 43.7% |
|  | 5 | 49.9% | 40.0% | 49.6% | 55.7% | 49.8% | 59.0% | 43.6% | 50.2% | 69.7% | 63.8% | 49.8% | 55.2% |
|  | 6 | 59.8% | 51.2% | 59.8% | 65.2% | 60.0% | 68.2% | 100.0% | 100.0% | 79.7% | 75.5% | 60.0% | 65.8% |
|  | 7 | 69.8% | 60.6% | 69.6% | 73.3% | 69.7% | 74.6% |  |  | 90.0% | 86.3% | 69.6% | 74.4% |
|  | 8 | 80.0% | 71.4% | 79.7% | 81.4% | 78.9% | 79.9% |  |  | 100.0% | 100.0% | 79.0% | 82.9% |
|  | 9 | 90.0% | 84.6% | 88.9% | 89.9% | 100.0% | 100.0% |  |  |  |  | 89.8% | 92.5% |
|  | 10 | 100.0% | 100.0% | 100.0% | 100.0% |  |  |  |  |  |  | 100.0% | 100.0% |
| Saguenay | 1 | 9.6% | 9.0% | 10.0% | 8.4% | 9.9% | 9.4% | 9.6% | 9.2% | 6.1% | 3.4% | 9.9% | 8.1% |
|  | 2 | 19.9% | 19.0% | 19.8% | 21.6% | 19.9% | 21.4% | 14.1% | 13.6% | 60.0% | 63.0% | 19.7% | 22.1% |
|  | 3 | 29.3% | 28.0% | 30.0% | 35.5% | 29.9% | 34.9% | 100.0% | 100.0% | 69.6% | 72.3% | 30.0% | 29.5% |
|  | 4 | 40.0% | 39.5% | 39.1% | 43.5% | 39.8% | 42.7% |  |  | 79.7% | 81.0% | 39.9% | 38.9% |
|  | 5 | 49.9% | 51.2% | 49.2% | 53.9% | 42.9% | 45.1% |  |  | 89.9% | 90.4% | 49.9% | 47.8% |
|  | 6 | 59.8% | 58.4% | 59.8% | 63.2% | 100.0% | 100.0% |  |  | 100.0% | 100.0% | 60.0% | 57.6% |
|  | 7 | 69.8% | 65.5% | 69.8% | 72.3% |  |  |  |  |  |  | 69.3% | 67.1% |
|  | 8 | 79.9% | 74.4% | 79.5% | 81.1% |  |  |  |  |  |  | 79.8% | 81.3% |
|  | 9 | 89.7% | 82.4% | 89.8% | 90.5% |  |  |  |  |  |  | 89.8% | 89.6% |
|  | 10 | 100.0% | 100.0% | 100.0% | 100.0% |  |  |  |  |  |  | 100.0% | 100.0% |
| Trois-Rivières | 1 | 9.9% | 10.7% | 9.9% | 11.3% | 9.3% | 13.1% | 9.8% | 11.5% | 6.6% | 5.4% | 9.8% | 8.4% |
|  | 2 | 19.8% | 20.4% | 19.8% | 22.8% | 19.7% | 22.9% | 19.4% | 23.3% | 48.9% | 45.5% | 19.7% | 18.9% |
|  | 3 | 29.7% | 29.0% | 30.0% | 31.9% | 30.0% | 33.3% | 28.5% | 32.4% | 59.5% | 56.2% | 29.9% | 30.6% |
|  | 4 | 40.0% | 38.1% | 39.6% | 40.6% | 39.6% | 45.0% | 30.1% | 33.6% | 70.0% | 68.4% | 40.0% | 38.7% |
|  | 5 | 49.9% | 46.4% | 49.0% | 48.7% | 48.6% | 54.0% | 100.0% | 100.0% | 79.9% | 78.9% | 49.4% | 50.0% |
|  | 6 | 59.8% | 58.9% | 59.2% | 60.1% | 59.9% | 63.1% |  |  | 89.9% | 89.6% | 59.3% | 59.3% |
|  | 7 | 69.8% | 67.4% | 69.5% | 71.4% | 61.4% | 64.7% |  |  | 100.0% | 100.0% | 69.4% | 67.7% |
|  | 8 | 79.7% | 77.1% | 79.7% | 79.9% | 100.0% | 100.0% |  |  |  |  | 79.8% | 78.0% |
|  | 9 | 89.8% | 87.4% | 89.8% | 88.9% |  |  |  |  |  |  | 89.5% | 87.0% |
|  | 10 | 100.0% | 100.0% | 100.0% | 100.0% |  |  |  |  |  |  | 100.0% | 100.0% |

\* Pop = cumulative proportion of population
\*\* Case = cumulative proportion of cases

**Figure S1.**
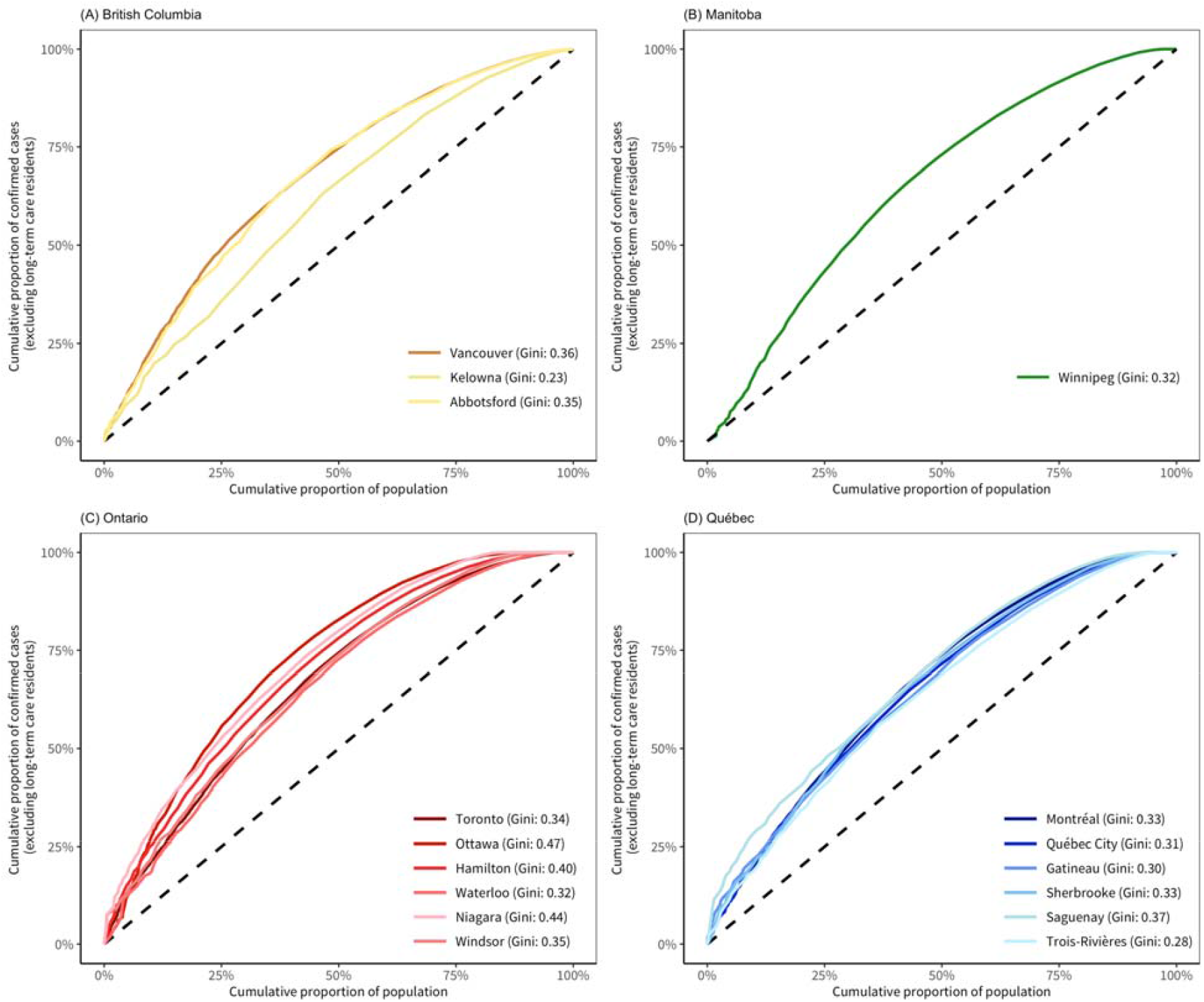
The Lorenz curves of COVID-19 confirmed cases (excluding long-term care residents) by proportion of population and the corresponding Gini coefficients. Panel A: Lorenz curves of census metropolitan areas (CMA) in British Columbia (Abbotsford-Mission is displayed as “Abbotsford”). Panel B: Lorenz curves of CMAs in Manitoba. Panel C: Lorenz curves of CMAs in Ontario (Ottawa-Gatineau (Ontario part) is displayed as “Ottawa”; Kitchener - Cambridge – Waterloo is displayed as “Waterloo”); St. Catharines–Niagara is displayed as “Niagara”). Panel D: Lorenz curves of CMAs in Québec (Ottawa-Gatineau (Québec part) is displayed as “Gatineau”). The population was ranked by the number of cases in each dissemination area (DA) from the highest to the lowest.

**Figure S2.**
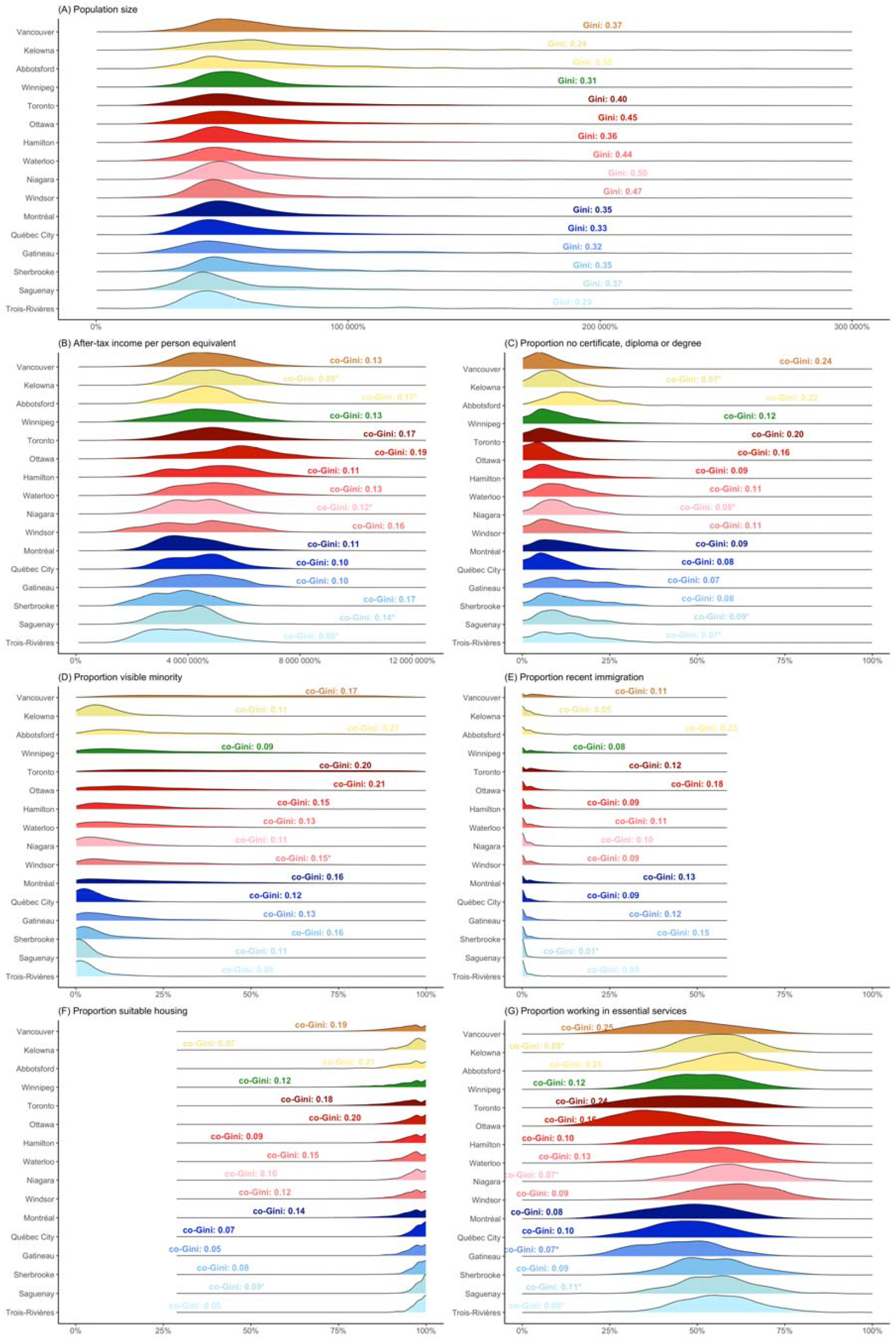
Distribution of the social determinants of health and the corresponding Gini (co-Gini) coefficients (excluding long-term care residents) of cumulative COVID-19 cases across census metropolitan areas (CMA). Panel A: population size. Panel B: After-tax income per person equivalent. Panel C: proportion population without certificate, diploma or degree deciles. Panel D: proportion visible minority. Panel E: proportion recent immigration. Panel F: proportion working in essential services. Panel G: proportion with suitable housing. Abbotsford-Mission is displayed as “Abbotsford”; Ottawa-Gatineau (Ontario part) is displayed as “Ottawa”; St. Catharines–Niagara is displayed as “Niagara”; Ottawa-Gatineau (Québec part) is displayed as “Gatineau”. Co-Gini coefficients followed by a “*” mark represent co-Gini coefficients of those Lorenz curves that went over and under the equality line.

**Figure S3.**
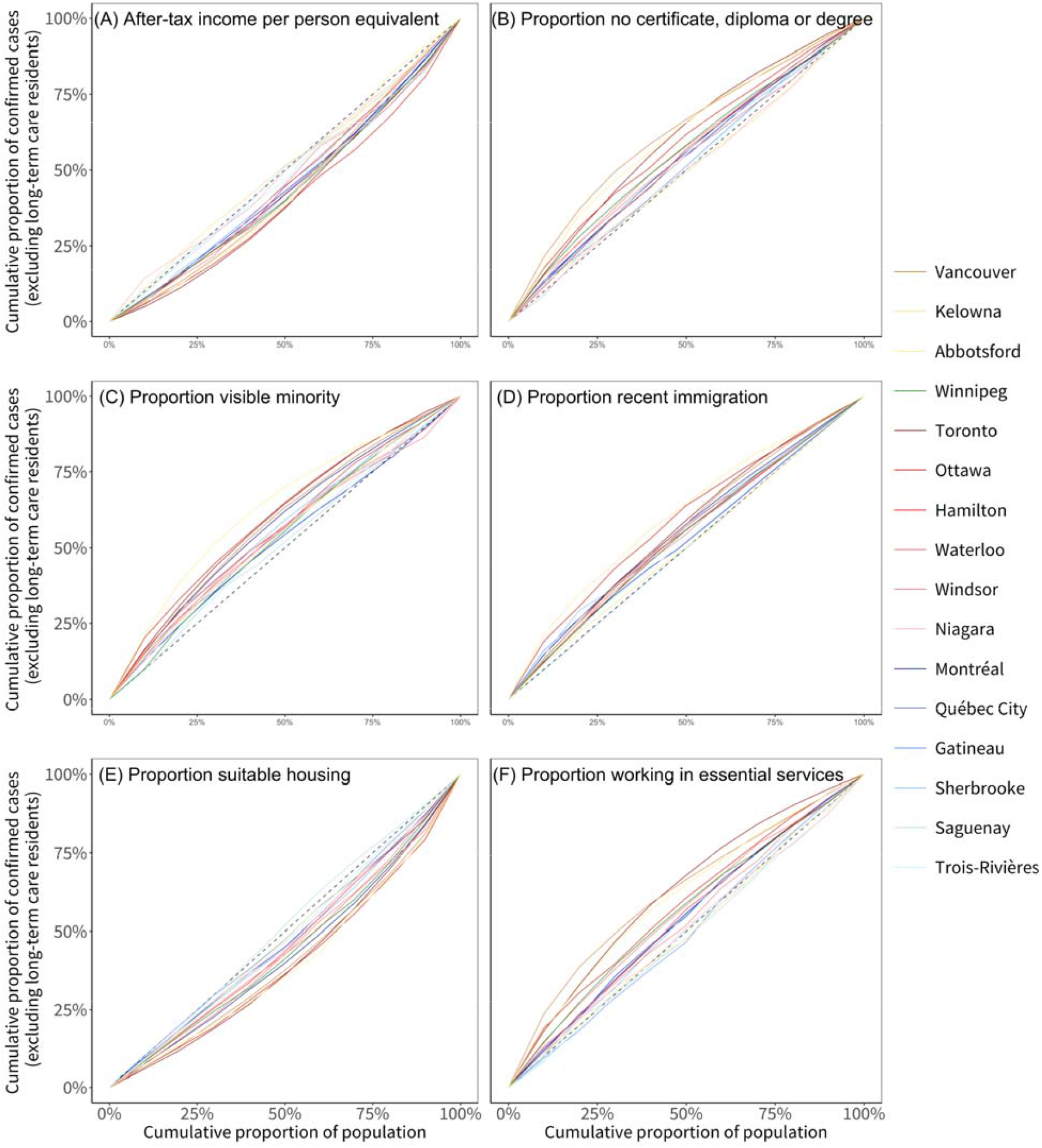
The concentration curves of COVID-19 confirmed cases (excluding long-term care residents) by social determinants. Panel A: after-tax income per-person equivalent deciles. Panel B: proportion population without certificate, diploma or degree deciles. Panel C: proportion visible minority deciles. Panel D: proportion recent immigration deciles. Panel E: proportion working in essential services deciles. Panel F: proportion with suitable housing deciles. Abbotsford-Mission is displayed as “Abbotsford”; Ottawa-Gatineau (Ontario part) is displayed as “Ottawa”; St. Catharines–Niagara is displayed as “Niagara”; Ottawa-Gatineau (Québec part) is displayed as “Gatineau”. All the variables were ranked from the highest value to the lowest.

## References

1. Coronavirus disease 2019 (COVID-19): Epidemiology update: Government of Canada; [Available from: https://health-infobase.canada.ca/covid-19/epidemiological-summary-covid-19-cases.html.

2. Public Health Infobase - Data on COVID-19 in Canada: Public Health Agency of Canada; [Available from: https://open.canada.ca/data/en/dataset/261c32ab-4cfd-4f81-9dea-7b64065690dc.

3. COVID-19 infections among healthcare workers and other people working in healthcare settings. Public Health Agency of Canada January 2021.

4. Solar O, Irwin A. A conceptual framework for action on the social determinants of health. Social Determinants of Health Discussion Paper 2 (Policy and Practice). Geneva: World Health Organization; 2010.

5. Crear-Perry J, Correa-de-Araujo R, Lewis Johnson T, McLemore MR, Neilson E, Wallace M. Social and Structural Determinants of Health Inequities in Maternal Health. J Womens Health (Larchmt). 2021;30(2):230–5.

6. Figueiredo AM, Figueiredo D, Gomes LB, Massuda A, Gil-Garcia E, Vianna RPT, et al. Social determinants of health and COVID-19 infection in Brazil: an analysis of the pandemic. Rev Bras Enferm. 2020;73(Suppl 2):e20200673.

7. Andersen LM, Harden SR, Sugg MM, Runkle JD, Lundquist TE. Analyzing the spatial determinants of local Covid-19 transmission in the United States. Sci Total Environ. 2021;754:142396.

8. Mollalo A, Vahedi B, Rivera KM. GIS-based spatial modeling of COVID-19 incidence rate in the continental United States. Sci Total Environ. 2020;728:138884.

9. Abrams EM, Szefler SJ. COVID-19 and the impact of social determinants of health. Lancet Respir Med. 2020;8(7):659–61.

10. Maroko AR, Nash D, Pavilonis BT. COVID-19 and inequity: a comparative spatial analysis of New York City and Chicago hot spots. J Urban Health. 2020;97(4):461–70.

11. Takian A, Kiani MM, Khanjankhani K. COVID-19 and the need to prioritize health equity and social determinants of health. Int J Public Health. 2020;65(5):521–3.

12. Ramirez IJ, Lee J. COVID-19 emergence and social and health determinants in Colorado: A rapid spatial analysis. Int J Environ Res Public Health. 2020;17(11).

13. Fielding-Miller RK, Sundaram ME, Brouwer K. Social determinants of COVID-19 mortality at the county level. PLOS ONE. 2020;15(10):e0240151.

14. Tai DBG, Shah A, Doubeni CA, Sia IG, Wieland ML. The disproportionate impact of COVID-19 on racial and ethnic minorities in the United States. Clin Infect Dis. 2021;72(4):703–6.

15. Cordes J, Castro MC. Spatial analysis of COVID-19 clusters and contextual factors in New York City. Spat Spatiotemporal Epidemiol. 2020;34:100355.

16. Census metropolitan area (CMA) and Census agglomeration (CA) Statistics Canada; [Available from: https://www150.statcan.gc.ca/n1/pub/92-195-x/2011001/geo/cma-rmr/cma-rmr-eng.htm.

17. Dictionary, Census of Population, 2016. Dissemination area (DA): Statistics Canada; 2016 [Available from: https://www12.statcan.gc.ca/census-recensement/2016/ref/dict/geo021-eng.cfm.

18. British Columbia Centre for Disease Control. COVID-19 surveillance case data, Public Health Reporting Data Warehouse. British Columbia Centre for Disease Control. (2020). 2021.

19. Postal Code Conversion File (PCCF), 2017. Statistics Canada Catalogue no. 92-154-X.

20. Census Profile, 2016 Census: Statistics Cnaada; [updated June 18, 2019. Available from: https://www12.statcan.gc.ca/census-recensement/2016/dp-pd/prof/index.cfm?Lang=E.

21. Statistics Canada. Postal Code Conversion File Plus (PCCF+) Version 7A & 7D. Ottawa: Minister of Industry; 2016.

22. Définition de cas de COVID-19 - Québec: Ministère de la Santé et des Services sociaux; [Available from: https://www.msss.gouv.qc.ca/professionnels/maladies-infectieuses/coronavirus-2019-ncov/.

23. The Lancet Respiratory M. COVID-19 transmission-up in the air. Lancet Respir Med. 2020;8(12):1159.

24. Sundaram ME, Calzavara A, Mishra S, Kustra R, Chan AK, Hamilton MA, et al. Individual and social determinants of SARS-CoV-2 testing and positivity in Ontario, Canada: a population-wide study. CMAJ. 2021.

25. Lee WC. Characterizing exposure-disease association in human populations using the Lorenz curve and Gini index. Stat Med. 1997;16(7):729–39.

26. Althaus CL, Turner KM, Schmid BV, Heijne JC, Kretzschmar M, Low N. Transmission of Chlamydia trachomatis through sexual partnerships: a comparison between three individual-based models and empirical data. J R Soc Interface. 2012;9(66):136–46.

27. R Core Team (2021). R: A language and environment for statistical computing. Vienna, Austria: R Foundation for Statistical Computing.

28. Thomas LJ, Huang P, Yin F, Luo XI, Almquist ZW, Hipp JR, et al. Spatial heterogeneity can lead to substantial local variations in COVID-19 timing and severity. Proceedings of the National Academy of Sciences. 2020;117(39):24180–7.

29. Mishra S, Stall NM, Ma H, Odutayo A, Kwong JC, Allen U. A vaccination strategy for ontario COVID-19 hotspots and essential workers: Science Table COVID-19 Advisory for Ontario; April 21, 2021 [Available from: https://covid19-sciencetable.ca/sciencebrief/a-vaccination-strategy-for-ontario-covid-19-hotspots-and-essential-workers/.

30. Simon L, John H. Some of B.C.’s COVID hot spots also have the lowest vaccination rates, data shows: Global News; [updated May 2021. Available from: https://globalnews.ca/news/7856112/bc-hot-spot-vaccination-rate-data-covid/.

31. Mody A, Pfeifauf K, Geng EH. Using Lorenz curves to measure racial inequities in COVID-19 testing. JAMA Network Open. 2021;4(1):e2032696–e.

32. Paton J. U.K. vaccination rates struggle in places worst-hit by Covid-19: Bloomberg Equality; March 30, 2021 [Available from: https://www.bloomberg.com/news/features/2021-03-30/britain-s-ethnic-vaccine-gap-risks-more-covid-19-cases-deaths-in-deprived-areas.

33. Questions and answers pertaining to employers and workers during the COVID-19 pandemic: Government of Québec; 2021 [updated May 27, 2021. Available from: https://www.quebec.ca/en/health/health-issues/a-z/2019-coronavirus/answers-questions-coronavirus-covid19/employers-workers-covid-19.

34. Jeyasundaram B. Community ambassadors are the link to Toronto’s unvaccinated populations: The Local; June 4, 2021 [Available from: https://thelocal.to/community-ambassadors-are-the-link-to-torontos-unvaccinated-populations/.

35. Chagla Z, Ma H, Sander B, Baral SD, Mishra S. Characterizing the disproportionate burden of SARS-CoV-2 variants of concern among essential workers in the Greater Toronto Area, Canada. medRxiv. 2021:2021.03.22.21254127.

36. Tirupathi R, Muradova V, Shekhar R, Salim SA, Al-Tawfiq JA, Palabindala V. COVID-19 disparity among racial and ethnic minorities in the US: A cross sectional analysis. Travel Medicine and Infectious Disease. 2020;38:101904.

37. Desmet K, Wacziarg R. Understanding spatial vriation in COVID-19 across the United States. J Urban Econ. 2021:103332.

38. Rozenfeld Y, Beam J, Maier H, Haggerson W, Boudreau K, Carlson J, et al. A model of disparities: risk factors associated with COVID-19 infection. Int J Equity Health. 2020;19(1):126.

39. Burstrom B, Tao W. Social determinants of health and inequalities in COVID-19. Eur J Public Health. 2020;30(4):617–8.

40. Palameta B. Low income among immigrants and visible minorities: Citeseer; 2004.

41. Shommu NS, Ahmed S, Rumana N, Barron GRS, McBrien KA, Turin TC. What is the scope of improving immigrant and ethnic minority healthcare using community navigators: A systematic scoping review. International Journal for Equity in Health. 2016;15(1):6.

42. Structural discrimination in COVID-19 workplace protections: Health Affairs Blog; May 29, 2020 [Available from: https://www.healthaffairs.org/do/10.1377/hblog20200522.280105/full/.

43. ‘It’s just a flu’: COVID prevention rules are often ignored on Toronto construction sites, workers tell the Star: Toronto Star; Feb. 16, 2021 [Available from: https://www.thestar.com/business/2021/02/16/its-just-a-flu-covid-prevention-rules-are-often-ignored-on-sites-worried-construction-workers-tell-the-star.html.

44. Thakur N, Lovinsky-Desir S, Bime C, Wisnivesky JP, Celedon JC. The structural and social determinants of the racial/ethnic disparities in the U.S. COVID-19 pandemic. What’s our role? Am J Respir Crit Care Med. 2020;202(7):943–9.

45. Berrigan D, Dodd K, Troiano RP, Krebs-Smith SM, Barbash RB. Patterns of health behavior in U.S. adults. Prev Med. 2003;36(5):615–23.

46. Mishra S, Ma H, Moloney G, Yiu KC, Darvin D, Landsman D, et al. Increasing concentration of COVID-19 by socioeconomic determinants and geography in Toronto, Canada: an observational study. medRxiv. 2021:2021.04.01.21254585.

47. Braveman P, Gottlieb L. The social determinants of health: it’s time to consider the causes of the causes. Public Health Rep. 2014;129 Suppl 2:19–31.

48. COVID-19 guidance for the health sector: Ontario Ministry of Health; [Available from: https://health.gov.on.ca/en/pro/programs/publichealth/coronavirus/2019_guidance.aspx#symptoms.

49. Testing for COVID-19: Government of Quebec; [Available from: https://www.quebec.ca/en/health/health-issues/a-z/2019-coronavirus/testing-for-covid-19/.

50. Godin A, Xia Y, Buckeridge DL, Mishra S, Douwes-Schultz D, Shen Y, et al. The role of case importation in explaining differences in early SARS-CoV-2 transmission dynamics in Canada-A mathematical modeling study of surveillance data. Int J Infect Dis. 2021;102:254–9.

